# Cardiovascular and non-cardiovascular risks among female breast cancer survivors in Japan: A matched cohort study

**DOI:** 10.1101/2024.10.10.24315278

**Authors:** Chitose Kawamura, Krishnan Bhaskaran, Takaaki Konishi, Yasuaki Sagara, Hiroko Bando, Tomohiro Shinozaki, Shuko Nojiri, Motohiko Adomi, Angel YS Wong, Nanako Tamiya, Masao Iwagami

**Author notes:** **Corresponding author;** Masao Iwagami, MD, MPH, MSc, PhD, Honorary Assistant Professor, Department of Non-Communicable Disease Epidemiology, London School of Hygiene and Tropical Medicine, Professor, Department of Health Services Research, Institute of Medicine, University of Tsukuba, 1-1-1 Tennodai, Tsukuba, Ibaraki, Japan, or.

## Abstract

**Background:** The number of breast cancer (BC) survivors has increased worldwide, but the landscape of their non-cancer disease risks remains unclear, especially among Asian women.

**Methods:** In the JMDC claims database, which covers company employees and their family members in Japan, women aged 18–74 years with and without an incident BC were matched in a 1:4 ratio for age and entry timing to the database between January 2005 and December 2019. The risks for six cardiovascular diseases (myocardial infarction, heart failure, atrial fibrillation/flutter, ischemic stroke, intracranial hemorrhage, and pulmonary embolism) and six non-cardiovascular diseases (major osteoporotic fractures, other fractures, gastrointestinal bleeding, urinary tract infection, infectious pneumonia, and anxiety/depression) were compared between the groups.

**Findings:** Comparing 24,017 BC survivors and 96,068 matched women (mean age, 50·5 years), the incidence rates of heart failure, atrial fibrillation/flutter, and all non-cardiovascular diseases were higher in the BC survivor group. The highest adjusted hazard ratio (HR) was noted for heart failure (4·09 [95% confidence interval 2·58–6·50]), followed by gastrointestinal bleeding (3·55 [3·10–4·06]), and anxiety/depression (3·06 [2·86–3·27]). The HRs in the first year were larger than those for 1–10 years for most outcomes, whereas the HRs for fracture outcomes were larger in the 1–10 years group.

**Interpretation:** BC survivors in Japan showed an increased risk of many non-cancer diseases compared to women without BC. Most risks increased more steeply during the first year following diagnosis, whereas the risk of fractures increased later.

**Funding:** Competitive research funding from Pfizer Health Research Foundation in Japan.

## Introduction

In 2020, breast cancer (BC) was the leading cause of cancer among women worldwide, with the highest incidence in most countries.^1^ Early detection and advancements in treatment strategies have increased survival rates for many patients with BC. For example, in the most recent data from Japan, the 5-year all-cause survival rates are 95·2% and 90·9% for patients with stage I and stage II, respectively.^2^

Accordingly, general health among BC survivors has become an increasing concern. As summarized in **Appendix pp 2–11**, American or European studies have suggested that BC survivors have an increased risk of several non-cancer diseases, such as heart failure and fractures.^3–6^ However, it remains controversial whether the risk of other diseases, such as myocardial infarction and stroke, also increases in BC survivors.^4,5^

Moreover, several questions remain unanswered. First, the aforementioned studies were conducted in American or European countries and included only a limited number of individuals of Asian descent (**Appendix p 11)**. Therefore, it remains unknown whether the risks of cardiovascular diseases are similarly heightened in Asian BC survivors. Second, regardless of ethnicity, there has been almost no data available on bleeding and infections comparing BC survivors with non-BC women, even though these are important diseases that require treatment and can sometimes be life-threatening. Third, only a small fraction of previous studies has considered BC treatment in their analyses, although the risks of individual cardiovascular and non-cardiovascular diseases may differ among patients receiving different treatment regimens.

To address these significant knowledge gaps, this study sought to investigate the risk of 12 major non-cancer diseases among BC survivors in comparison to age-matched women. The diseases examined include six cardiovascular conditions (myocardial infarction, heart failure, atrial fibrillation/flutter, ischemic stroke, intracranial hemorrhage, and pulmonary embolism) and six major non-cardiovascular conditions (osteoporotic fractures, other fractures, gastrointestinal bleeding, infectious pneumonia, urinary tract infections, and anxiety/depression). This analysis utilized a large, population-based claims database from Japan.

## Methods

### Study design

We conducted a matched cohort study using data from the JMDC claims database from January 2005 to June 2022.

### Data source

The JMDC Co. has obtained individual medical claims and annual health checkup data from participating health insurance associations for large- and medium-sized company employees and their family members aged <75 years.^7^ To date, several clinical and health-service studies have been conducted using the JMDC database. ^8–10^ Since 2005, the number of participating health insurance associations, and therefore the number of insured individuals in the JMDC database, has consistently increased, reaching a cumulative total of >14 million people when we obtained the studied data at the end of 2022. In addition to demographics and eligibility information, the JMDC database includes monthly data on inpatient and outpatient diagnoses recorded based on the International Classification of Diseases and Related Health Problems, 10th Revision (ICD-10) codes, prescribed and dispensed medications (in both hospitals and clinics) coded using the World Health Organization Anatomical Therapeutic Chemical classifications,^11^ European Pharmaceutical Market Research Association codes, and procedures such as surgery, using original Japanese codes.

This study was approved by the Ethics Committee of the University of Tsukuba, Ibaraki, Japan (approval number 1839). The requirement for consent from individual participants was waived because the data were anonymized.

### Study participants

The exposed group, or BC survivors, included women with the incident BC, defined as a new diagnosis of BC (ICD-10 C50) between database inception (January 2005) and December 2019, who also received BC surgery (Japanese original procedure codes K476.1-K476.9) within 1 year of diagnosis. We selected December 2019 because the COVID-19 pandemic could have delayed BC diagnosis after January 2020 in Japan and led to misclassification of BC status. To ensure that the BC diagnosis was new, we excluded patients with possibly prevalent BC, defined as a diagnosis within 3 months of entering the JMDC database. We also excluded patients indicating metastatic BC, recurrent BC, sarcoma, or malignant phyllodes tumors using the Japanese original diagnosis codes, as they have different clinical courses from primary BC survivors. Moreover, we excluded patients with BC whose diagnosis and JMDC withdrawal occurred in the same month because their follow-up lengths were almost non-existent. The month of the first BC diagnosis was referred to as the “index month” to indicate the start of follow-up.

For each BC survivor, we randomly selected four women with no history of BC at the index month, with the same age (±1 year) and the same timing (year and month) of entry to the JMDC database to ensure the same lengths of covariate definitions between the groups. The sampling was conducted without replacement, meaning that a woman could be selected for the matched cohort only one time.

### Outcomes

We pre-defined algorithms to define the 12 outcomes, using ICD-10 codes; hospitalization; medical procedures, such as surgery, ablation, and orthopedic cast; and medication (**Appendix pp 12–14**) in reference to previous studies using the JMDC databases and validation studies using Japanese claims data.^12^ We examined six major cardiovascular diseases, including myocardial infarction (inpatient diagnosis and cardiac catheter or coronary artery bypass grafting), heart failure (inpatient diagnosis and cardiac echocardiography and diuretics or inotropic drugs), atrial fibrillation/atrial flutter (inpatient/outpatient diagnosis and catheter ablation or maze operation or antiarrhythmic drugs or anticoagulants), ischemic stroke (inpatient diagnosis and catheter or surgical thrombus retrieval or thrombectomy or specific antithrombotic drugs including tissue plasminogen activator, antiplatelet drugs, or specific anticoagulants), intracranial hemorrhage (inpatient diagnosis and specific surgery or antihypertensive or specific nitrate drugs), and pulmonary embolism (inpatient diagnosis and pulmonary embolectomy or inferior vena cava filter placement or specific antithrombotic drugs) and six major non-cardiovascular diseases including major osteoporotic fractures (thoracolumbar and pelvic fractures, proximal humerus fractures, distal radius fractures, and femoral neck fractures; inpatient/outpatient diagnosis and orthopedic surgery or orthopedic procedures or orthopedic cast), other fractures (all fractures except for major osteoporotic fractures; inpatient/outpatient diagnosis and orthopedic surgery or orthopedic procedures or orthopedic cast), gastrointestinal bleeding (inpatient/outpatient diagnosis and endoscopic procedure), infectious pneumonia (inpatient/outpatient diagnosis and antibiotics or antifungal drug or antimycobacterial drugs or antiviral drugs), urinary tract infection (inpatient/outpatient diagnosis and antibiotics or antifungal drugs), and anxiety/depression (inpatient/outpatient diagnosis and antianxiety drugs or antidepressants). Anxiety and depression were grouped together as they are often recorded together or misclassified for reimbursement purposes in the Japanese claims system.

### Covariates

Potential confounding factors between BC survivorship and the studied outcomes included body mass index (BMI) (classified as <18·5 kg/m², 18·5–24·9 kg/m², 25·0–29·9 kg/m², or ≥30·0 kg/m²), smoking history (non-smoker/ex-smoker or current smoker), drinking habits (none, sometimes, or every day), and common lifestyle-associated diseases, including hypertension, diabetes, and dyslipidemia. In both groups, information on BMI, smoking history, and drinking habits was obtained from the most recent annual health checkup data at or before the index month. Hypertension, diabetes, and dyslipidemia were defined based on any history of prescriptions for these conditions from JMDC enrollment to the index month. Additionally, for fracture outcomes, we included patients treated for osteoporosis based on any history of prescriptions for osteoporosis before the index month.

### Statistical analysis

We first summarized the baseline characteristics in each group and compared these between the groups using t-tests for age and chi-square tests for BMI, smoking status, drinking habits, comorbidities, and previous histories of each outcome.

Subsequently, we calculated the crude incidence rate for each outcome in each group and the crude difference and ratio between the groups. For each outcome, both groups were followed-up from the index month to the earliest incidence of that outcome, withdrawal from the JMDC database (which suggests that the patient lost insurance due to death or ceasing to be an employee, or that the health insurance association no longer contributed to the JMDC database), June 2022, or 10 years from the index month. Additionally, if an individual in the matched cohort group was subsequently diagnosed with BC, she was censored in the month of the BC diagnosis because she would be included in the BC survivor group from that point forward.

For each outcome, patients with a record of that outcome in the same month as the index month or earlier were considered to have a past medical history and were excluded from that analysis. We plotted Kaplan–Meier survival curves for each outcome.

As a sensitivity analysis, both groups were followed-up from the month of the first BC surgery (and the same month for the matched women without BC), instead of the index month (i.e., the month of the first BC diagnosis) in the main analysis.

We then estimated adjusted hazard ratios (HRs) using stratified Cox regression models to account for the matching: model 1 without additional adjustment; model 2 adjusted for hypertension, diabetes, and dyslipidemia, in addition to osteoporosis for the major osteoporotic fractures and other fracture outcomes; model 3 further adjusted for BMI, smoking history, and drinking habits as a complete case analysis; and model 4 included multiple imputation for BMI, smoking history, and drinking habits using information from other variables (i.e., exposure status, hypertension, diabetes, dyslipidemia, osteoporosis [only in the analysis of major osteoporotic fractures and other fractures], and outcome status in each analysis).^13^

Further, after observing the Kaplan–Meier survival curves and corresponding cumulative incidence curves plotted on a logarithmic scale (as shown later), we decided to estimate and report the incidence rates and HRs separately for <1 year and 1–10 years. This was because several outcomes showed evidently different trends between these two periods, consistent with the fact that most multidisciplinary treatments for BC (e.g., chemotherapy), except for hormone treatment, are completed within 1 year of diagnosis.

As part of an additional analysis, we estimated adjusted HRs by treatment regimens during the first year following the index month, contingent upon having a sufficient outcome number. We categorized patients with BC into four chemotherapy groups: anthracycline, taxane, anthracycline and taxane, and no anthracycline or taxane (for heart failure outcomes, we utilized human epidermal growth factor receptor 2 (HER2)-targeted therapy in place of taxane). Similarly, by hormone therapy, patients with BC were divided into three groups: tamoxifen, aromatase inhibitors, and neither tamoxifen nor aromatase inhibitors. Patients receiving both tamoxifen and aromatase inhibitors (n=348) were excluded from this analysis. Patients experiencing any event of interest or loss to follow-up within the first year were also excluded. For this analysis, we employed simple Cox regression rather than stratified Cox regression to compare BC patients receiving each treatment with non-BC women, adjusting for age, year of diagnosis, comorbidities (including osteoporosis for fracture outcomes), axillary dissection, radiotherapy, as well as the administration of anthracyclines and taxanes (using HER2-targeted therapy instead of taxanes for heart failure) when analyzed by hormone therapy, and tamoxifen and aromatase inhibitors when analyzed by chemotherapy.

All analyses were performed using STATA version 15.1 (Stata Corp, College Station, TX, USA).

## Results

Between January 2005 and December 2019, we included a total of 24,017 BC survivors (mean age, 50·5 years) and 96,068 women with no history of BC (mean age, 50·5 years), of whom 822 were later diagnosed with BC and therefore censored (**Figure 1**). **Table 1** presents the baseline characteristics of the groups. BC survivors showed a higher prevalence of hypertension and dyslipidemia, as well as a greater number of previous outcomes (heart failure, major osteoporotic fractures, other fractures, gastrointestinal bleeding, infectious pneumonia, urinary tract infection, and anxiety/depression) compared to the matched cohort group. The treatment details of the BC survivors during the first year are presented in **Appendix p 15**.

**Figure 1.**
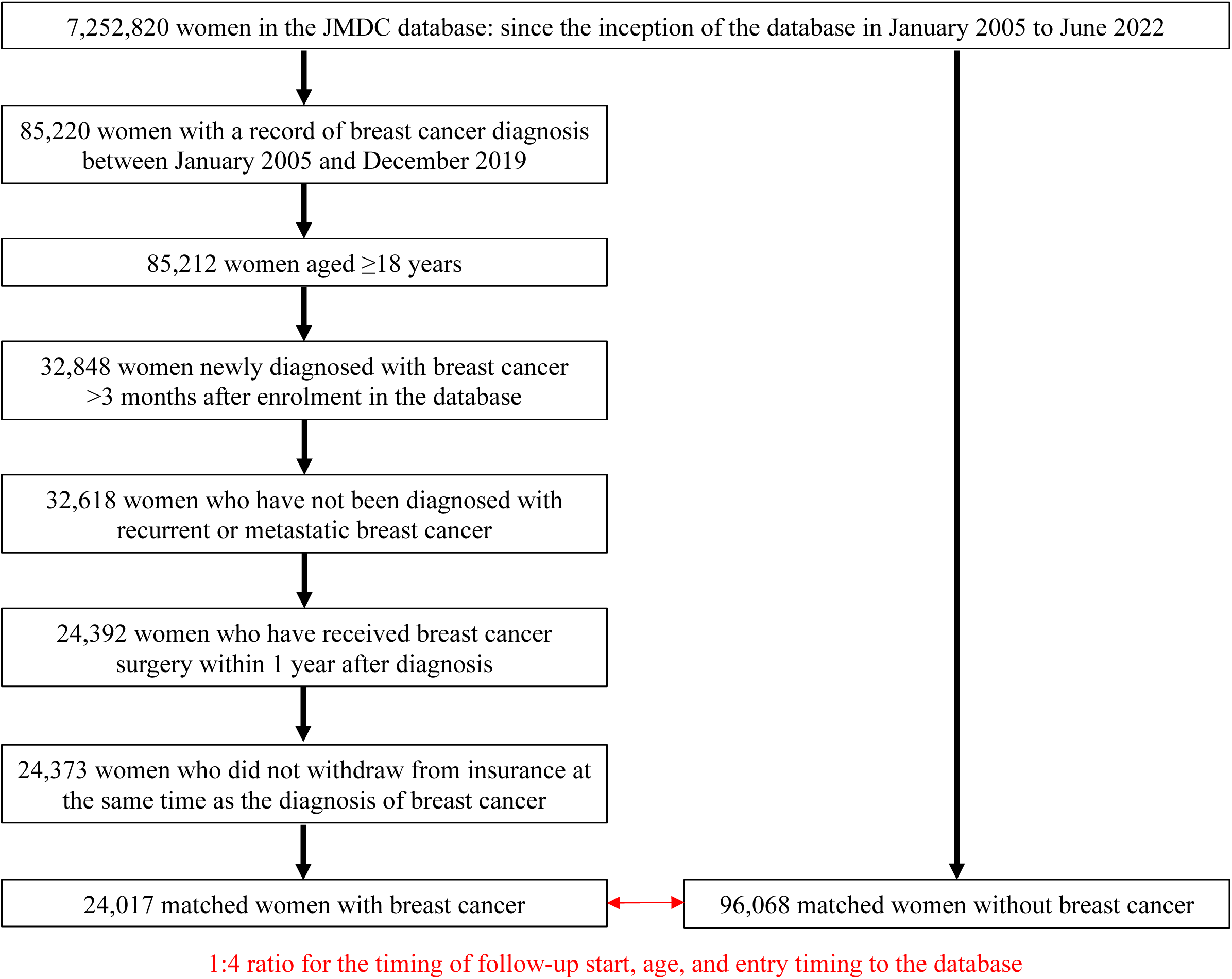
Flow chart of matched cohort of women with and without breast cancer.

**Table 1.** Baseline characteristics of matched women with and without breast cancer.

|  | Patients with<br>breast cancer<br>N=24,017<br>n (%) | Women without<br>breast cancer<br>N=96,068<br>n (%) | P-value |
| --- | --- | --- | --- |
| Age (years), mean (SD) | 50.5 (8.7) | 50.5 (8.7) | >0.999 |
| Body mass index |  |  | <0.001 |
| Underweight (<18.5 kg/m<sup>2</sup>) | 2,044 (8.5) | 8,118 (8.5) |  |
| Normal weight (18.5–24.9 kg/m<sup>2</sup>) | 11,813 (49.2) | 42,638 (44.4) |  |
| Overweight (25–29.9 kg/m<sup>2</sup>) | 2,532 (10.5) | 8,901 (9.3) |  |
| Obese (≥30 kg/m<sup>2</sup>) | 699 (2.9) | 2,742 (2.9) |  |
| Missing | 6,929 (28.9) | 33,669 (35.1) |  |
| Smoking status |  |  | <0.001 |
| Non-smoker/ex-smoker | 15,262 (63.6) | 54,972 (57.2) |  |
| Current smoker | 1,625 (6.8) | 6,727 (7.0) |  |
| Missing | 7,130 (29.7) | 34,369 (35.8) |  |
| Drinking habits |  |  | <0.001 |
| None | 8,793 (36.6) | 33,411 (34.8) |  |
| Sometimes | 5,078 (21.1) | 17,893 (18.6) |  |
| Every day | 2,406 (10.0) | 7,901 (8.2) |  |
| Missing | 7,740 (32.2) | 36,863 (38.4) |  |
| Comorbidities |  |  |  |
| Diabetes | 786 (3.3) | 2,933 (3.1) | 0.079 |
| Dyslipidemia | 2,876 (12.0) | 10,663 (11.1) | <0.001 |
| Hypertension | 3,157 (13.1) | 11,539 (12.0) | <0.001 |
| Osteoporosis | 669 (2.8) | 3,038 (3.2) | 0.002 |
| Past medical history |  |  |  |
| Myocardial infarction | 6 (0.02) | 9 (0.01) | 0.053 |
| Heart failure | 28 (0.12) | 69 (0.07) | 0.029 |
| Atrial fibrillation/flutter | 79 (0.33) | 277 (0.29) | 0.301 |
| Ischemic stroke | 18 (0.07) | 125 (0.13) | 0.027 |
| Intracranial hemorrhage | 14 (0.06) | 86 (0.09) | 0.133 |
| Pulmonary embolism | 11 (0.05) | 24 (0.02) | 0.091 |
| Major osteoporotic fractures | 175 (0.73) | 530 (0.55) | 0.001 |
| Other fractures | 630 (2.6) | 1,626 (1.7) | <0.001 |
| Gastrointestinal bleeding | 341 (1.4) | 652 (0.68) | <0.001 |
| Infectious pneumonia | 755 (3.1) | 2,680 (2.8) | 0.003 |
| Urinary tract infection | 2,900(12.1) | 10,612 (11.1) | <0.001 |
| Anxiety/depression | 1,760 (7.3) | 5,777 (6.0) | <0.001 |

SD = standard deviation.

**Table 2** shows the crude incidence rate for each outcome in each group, along with the differences and ratios between the groups. The incidence rates of heart failure, atrial fibrillation/flutter, and all non-cardiovascular diseases were higher in the BC survivors than in the matched cohort group, with the lower limit of the 95% confidence interval (CI) for their difference exceeding 0. The highest adjusted HR in the last model (model 4) was noted for heart failure (4·09, 95% CI 2·58–6·50), followed by gastrointestinal bleeding (3·55, 95% CI 3·10–4·06) and anxiety/depression (3·06, 95% CI 2·86–3·27).

**Table 2.**
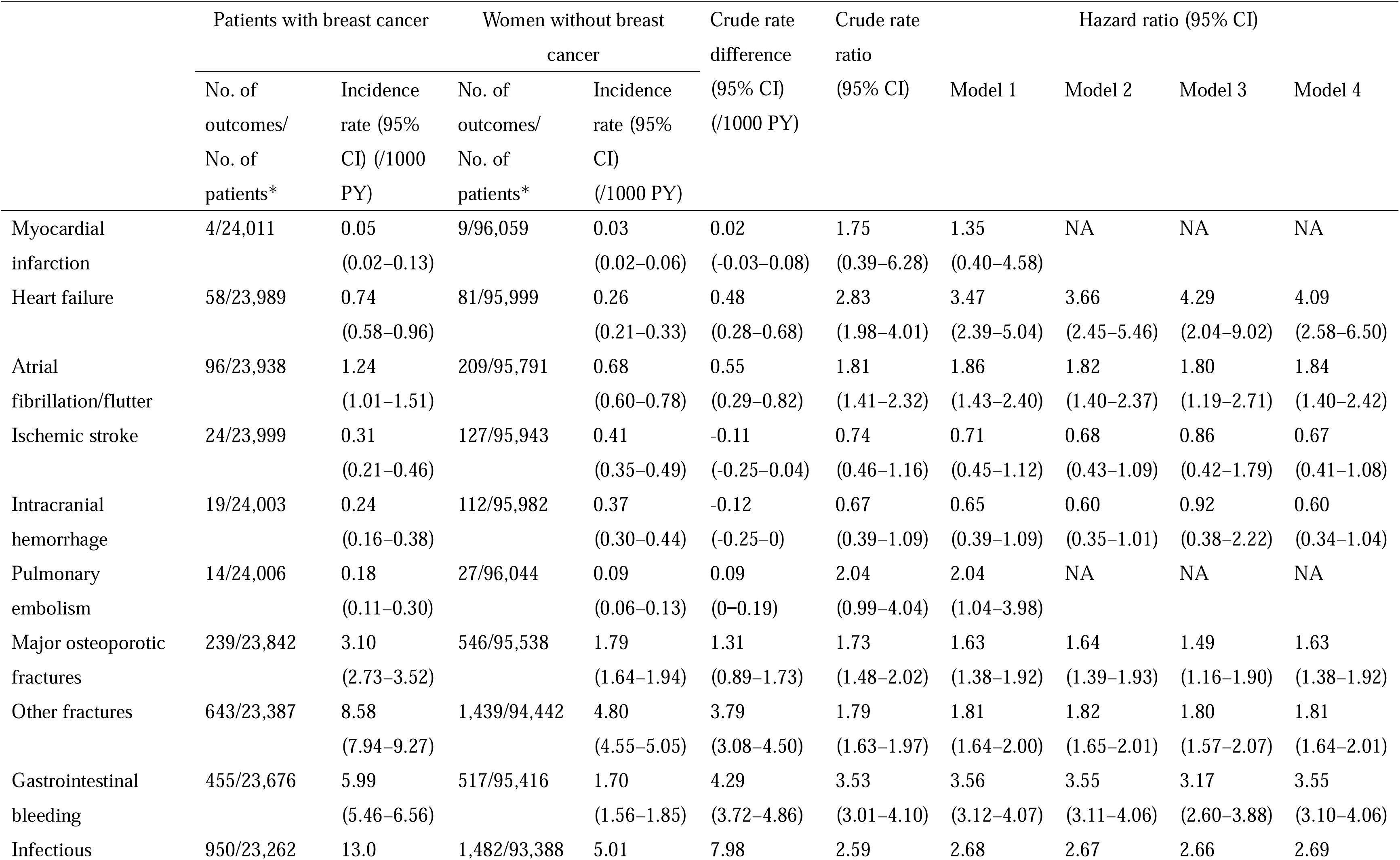
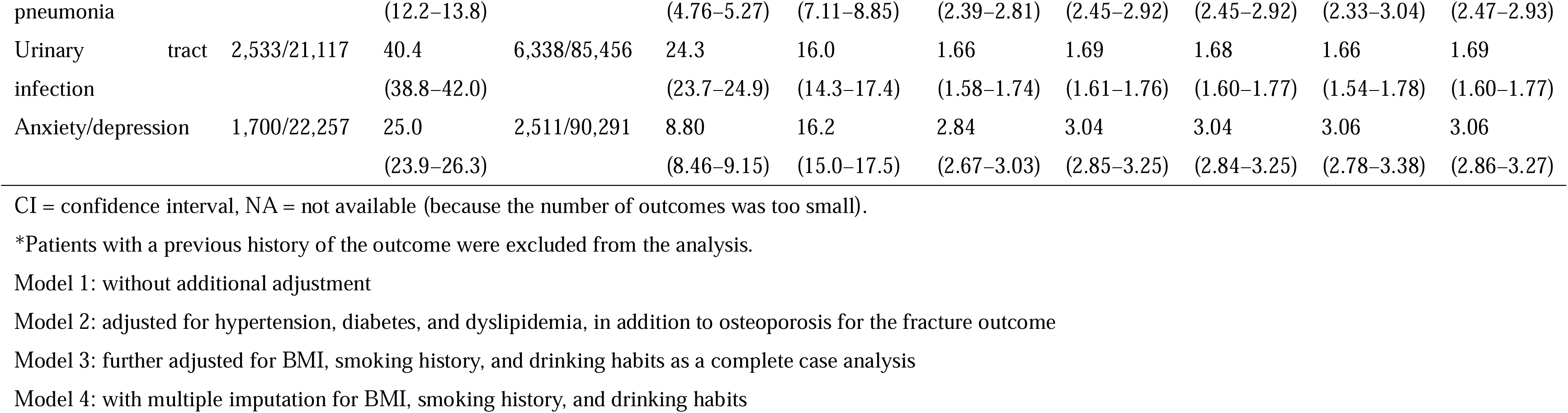
Incidence rate of each outcome in each group and its difference and ratio between the groups with and without breast cancer.

**Figure 2** shows the Kaplan–Meier curves of the cumulative incidence for each outcome, while **Appendix pp 16–17** presents the corresponding cumulative incidence curves plotted on a logarithmic scale. The incidences of all outcomes in the matched cohort group consistently increased over time. In contrast, the incidences of some outcomes, such as infectious pneumonia and anxiety/depression, increased more steeply during the first year of follow-up, whereas the incidence of major osteoporotic fractures and other fractures increased more steeply later in the follow-up period.

**Figure 2.**
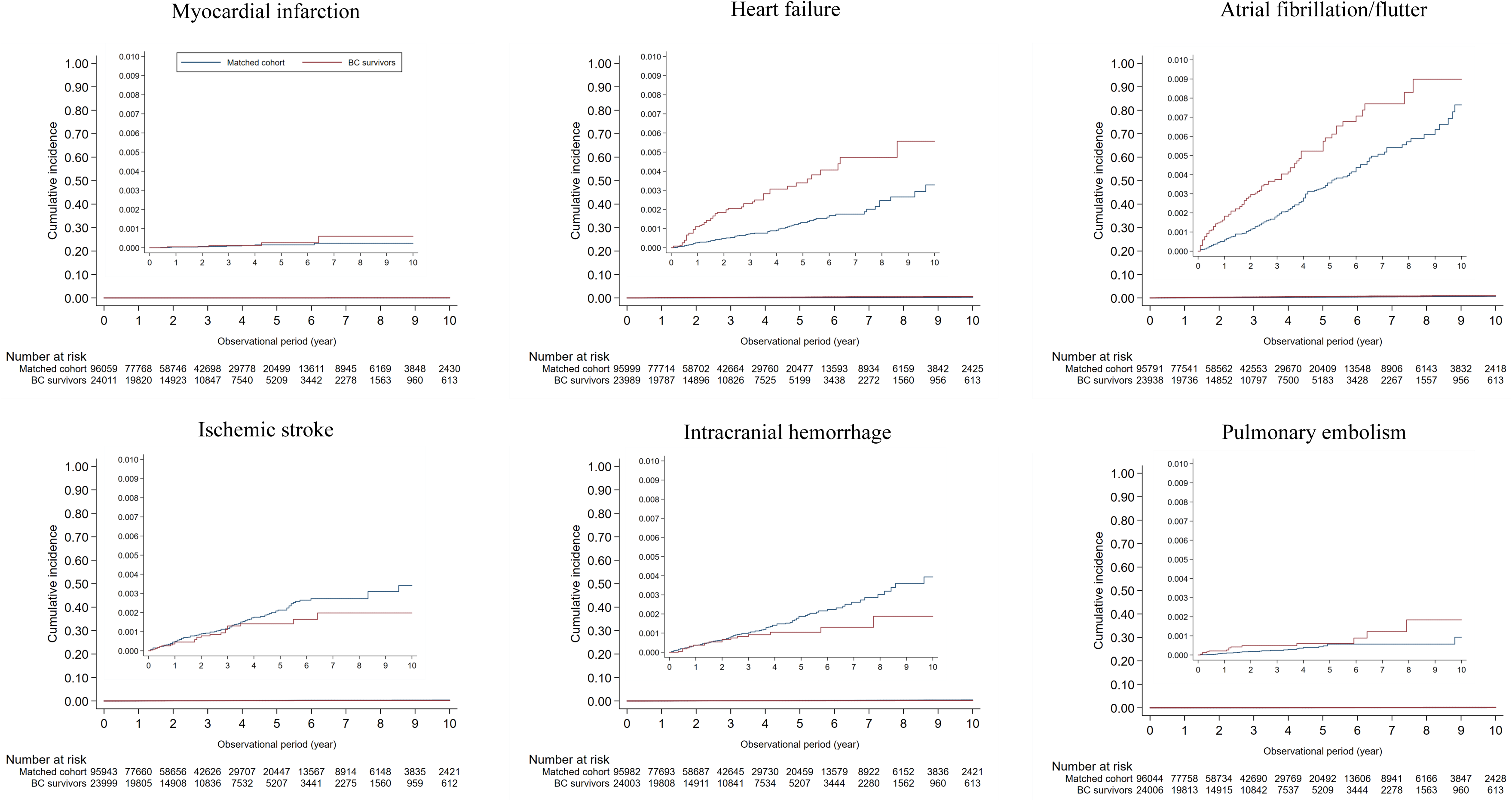
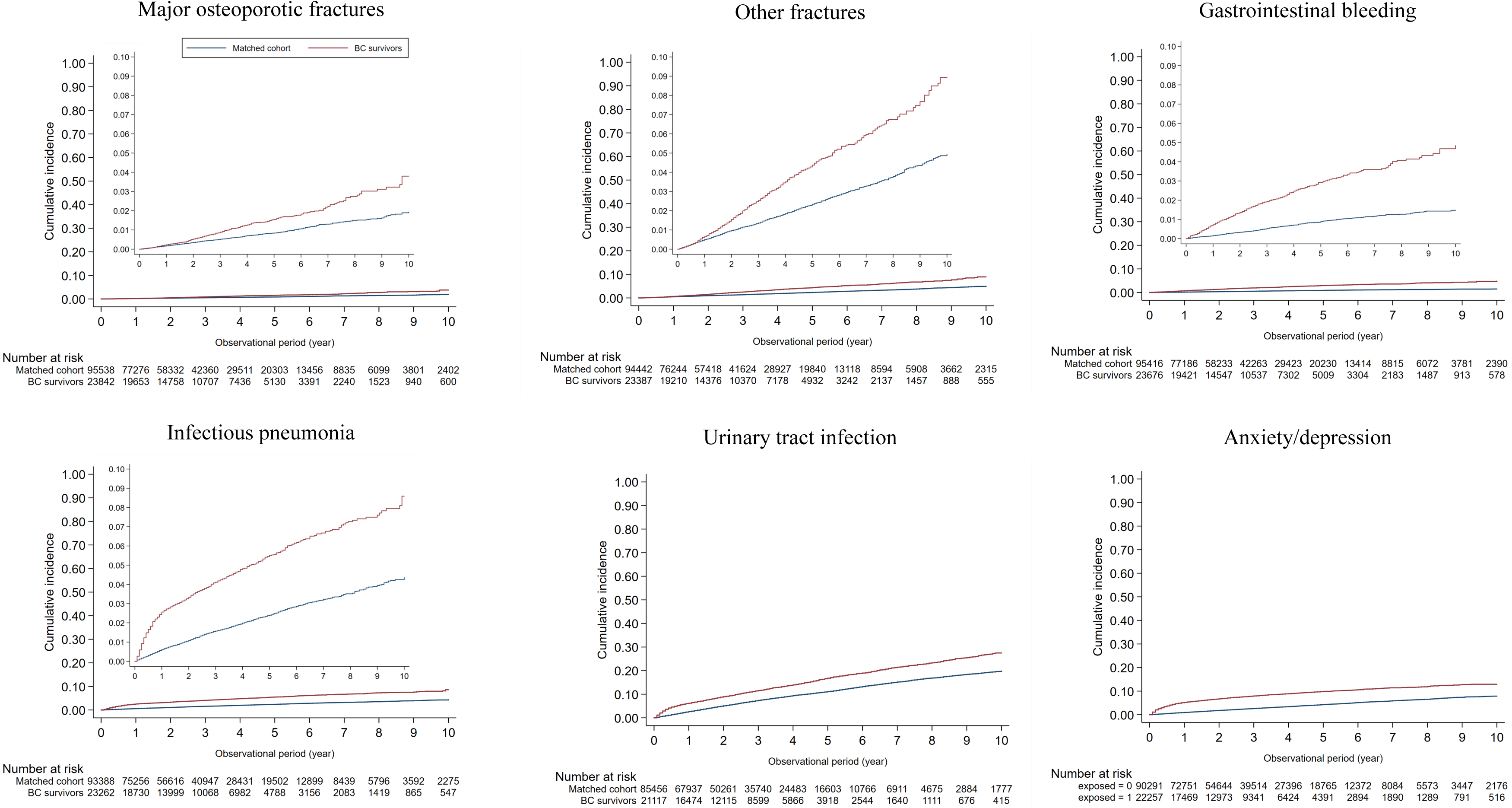
Kaplan–Meier curves of the cumulative incidence among women with and without breast cancer for each outcome.

Results of sensitivity analysis starting the follow-up from the month of the first BC surgery (and the same month for the matched women without BC) were very similar to those in the main analysis (**Appendix pp 18–19**).

**Appendix pp 20–23** presents the results of survival analysis separately for <1 year and 1–10 years. The adjusted HRs in the last model (model 4), excluding those for myocardial infarction and pulmonary embolism (which could not be obtained because of the small number of outcomes), are summarized graphically in **Figure 3**. The HRs for <1 year were larger than those for 1–10 years from the BC diagnosis for most outcomes, especially anxiety/depression (<1 year 5·99 [95% CI 5·42–6·61], and 1–10 years 1·51 [95% CI 1·37–1·67]). However, the HR was larger for 1–10 years than for <1 year for major osteoporotic fractures (<1 year 1·30 [95% CI 0·92–1·83], and 1–10 years 1·78 [95% CI 1·46–2·17]) and other fractures (<1 year 1·28 [95% CI 1·05–1·56], and 1–10 years 2·09 [95% CI 1·86–2·34]).

**Figure 3.**
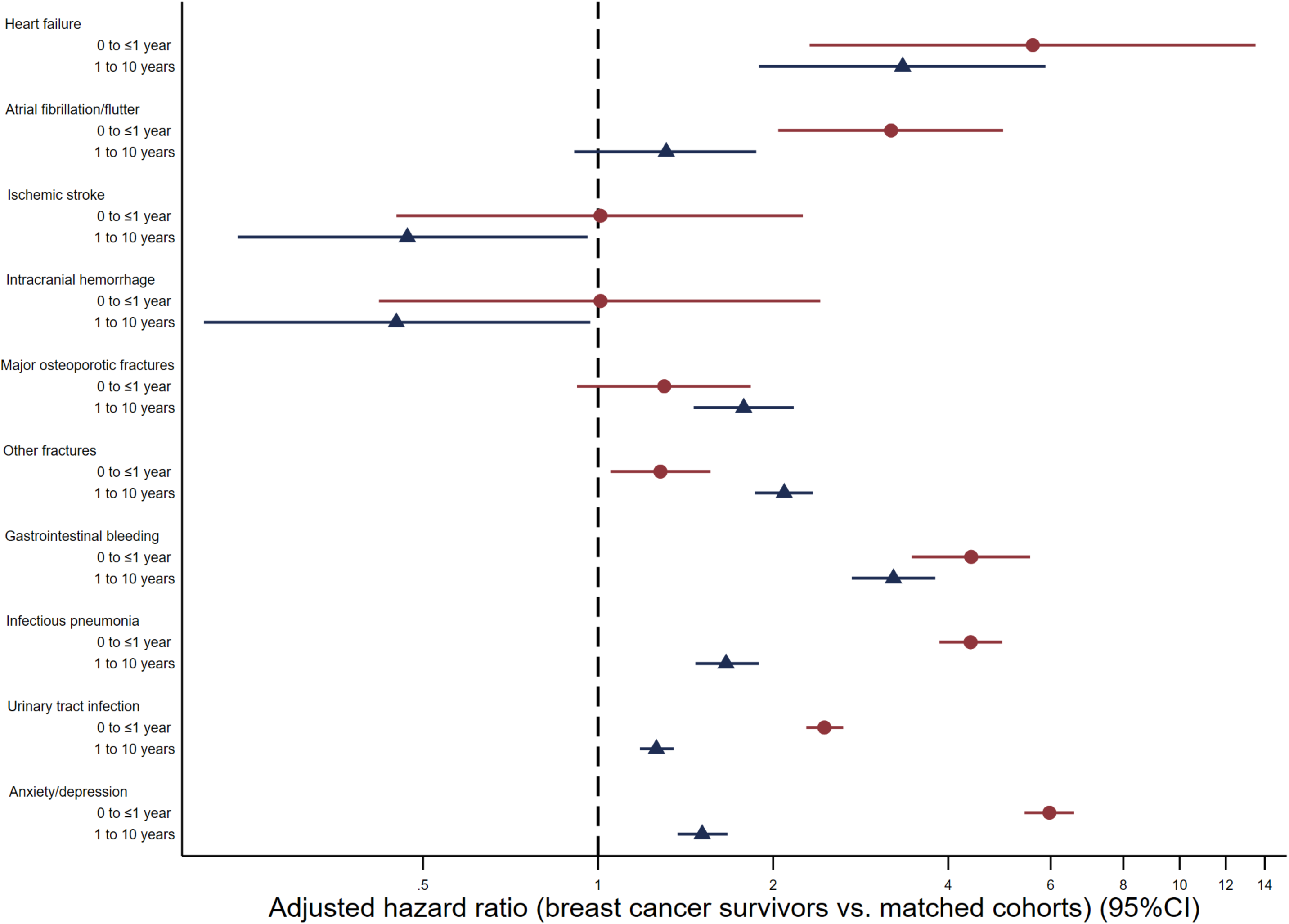
Adjusted hazard ratios between women with and without breast cancer for each outcome during <1 year and 1–10 years from the index month. CI = confidence interval, BMI = body mass index. Note: The adjusted hazard ratios (95% CIs) are from the last model (model 4) adjusted for hypertension, diabetes, dyslipidemia, osteoporosis (only for the fracture outcome), BMI, smoking history, and drinking habits, with multiple imputation for BMI, smoking history, and drinking habits (**Appendix pp 20–23**).

The additional analysis by treatment regimens was conducted for heart failure, atrial fibrillation/flutter, and all non-cardiovascular diseases, which showed a sufficient number of outcomes (**Appendix pp 24–31**). For chemotherapy, the point estimates of adjusted HRs for major osteoporotic fractures, other fractures, gastrointestinal bleeding, infectious pneumonia, and anxiety/depression in the anthracycline and taxane group, as well as that for heart failure in the anthracycline and HER2-targeted therapy group, were higher than in the other groups, although their 95% CIs were overlapping. For hormone therapy, the point estimates of adjusted HRs for major osteoporotic fractures and gastrointestinal bleeding were higher in the aromatase inhibitor group than in the other groups, although their 95% CIs were overlapping.

## Discussion

Using a large Japanese claims database that includes over 24,000 women with BC, we illustrated the landscape of non-cancer risks among Japanese female BC survivors compared with the general population of women of the same age. Japanese BC survivors showed an increased risk of heart failure, atrial fibrillation/flutter, major osteoporotic fractures, other fractures, gastrointestinal bleeding, infectious pneumonia, urinary tract infections, and anxiety/depression compared to women without BC. The HRs at less than 1 year were higher than those at 1–10 years of follow-up for most outcomes, especially anxiety/depression. However, the opposite trend was observed for fractures. We also found that the risk of certain outcomes could differ according to various treatment regimens for BC, although the analysis appeared to be underpowered even with this large database.

Previous American and European studies have suggested that the overall risk of cardiovascular diseases among patients with BC is high; ^14^ however, there is both agreement (e.g., heart failure) and controversy (e.g., stroke) regarding the risk associated with each type of cardiovascular disease. Consistent with previous studies, BC survivors in Japan were more likely to develop heart failure than age-matched women without BC, with even higher ratios than those reported in American and European studies ^3,4,15–17^ BC survivors in Japan were more likely to develop heart failure than age-matched women without BC, with even higher ratios than those reported in the previous studies. Although previous studies showed differing risks for other cardiovascular diseases,^3,4,15–19^ our study indicated a higher risk of atrial fibrillation/atrial flutter but no strong evidence for higher risk of myocardial infarction, ischemic stroke, intracranial hemorrhage, or pulmonary embolism. Differences in results between studies may be attributable to variations in the age distribution of the study population, the baseline risk of each outcome by country or race, and the stage of BC at detection. The overall study population was younger (mean age of 50·5 years) than most previous studies (**Appendix pp 2–10**), mainly because the database does not include individuals aged 75 years and older, all of whom are covered by the Late Elders’ Health Insurance in Japan. It is possible that the Japanese in this population were less likely to experience cardiovascular events than individuals of similar age in other countries. BC and cardiovascular diseases have several overlapping risk factors.^20,21^ Although cardiovascular diseases can be identified early, effectively managed, and in many cases prevented,^22^ cardiovascular disease screening and management do not typically feature in routine long-term BC care.^23^ It is important to ensure that regular checks for treatment-specific adverse events are performed (e.g., echocardiography to evaluate cardiac function in patients treated with anthracycline or HER2-targeted treatments) and that patients are encouraged to undergo medical screening (e.g., electrocardiogram or measurement of blood pressure), which is not usually performed routinely as part of BC care. Prevention and early treatment (e.g., statins, angiotensin-converting enzyme inhibitors/angiotensin receptor blockers) are also possible.

Many studies have indicated that BC survivors have more osteoporotic fractures, both pre-and post-menopausal (**Appendix pp 2–11**). Osteoporosis due to aromatase inhibitors and chemotherapy-induced ovarian failure is an important risk factor for fractures in BC survivors.^24,25^ Although the participants in this study were relatively young, the risk for both osteoporotic and other fractures was higher among BC survivors, which is consistent with results from the previous large population-based studies. BC patients receive hormone therapy for 5 to 10 years. Although the number at risk after 5 years is not large in our study, it is conceivable that the risk could increase as more cases are observed over longer periods. It is important to consider whether bone density tests are performed regularly on those at higher risk (e.g., due to aromatase inhibitors or early menopause from chemotherapy) and whether patients with reduced bone density receive appropriate treatment, such as bisphosphonates and parathyroid hormone. Moreover, it has been reported that osteoporotic fractures in BC patients are not solely due to decreased bone density,^26^ and it is important for physicians not only to check bone density but also to encourage patients to quit smoking, eat a healthy diet, and adopt exercise habits, while informing them of their fracture risk to prevent osteoporotic fractures. For other fractures (e.g., rib fractures), as with osteoporotic fractures, it might be important to assess bone density, educate patients, and inform them of their fracture risk.

BC survivors have higher risks of mental health diseases, such as depression, anxiety, and stress-related disorders than non-BC women,^27^ and these risks are highest from diagnosis to approximately 1 year after diagnosis.^5,28,29^ Treatment with chemotherapy is also known to be a risk factor for mental health diseases.^5,28^ In our study, BC survivors had a higher risk of anxiety/depression than non-BC women, with the highest HR occurring within 1 year of diagnosis. One year after diagnosis, the HR (for the first incidence of anxiety/depression in the database) declined but remained higher than that of non-BC women. It is important for healthcare providers to keep in mind that BC patients are at high risk for anxiety or depression, even 1 year after treatment, and to consider psychosocial interventions as needed. To the best of our knowledge, this is the first study to demonstrate that patients with BC have a higher risk of developing gastrointestinal bleeding and infections. Notably, the risk was high not only within the first year after the diagnosis of BC when treatment was intensive (e.g., chemotherapy) but also beyond 1 year. The increased risks of infections may suggest that BC survivors have decreased immunity associated with BC and/or side effects of BC treatments. Possible mechanisms for increased gastrointestinal bleeding include stress from BC, damage to the vessel wall due to side effects of BC treatment, the use of pain medications such as non-steroidal anti-inflammatory drugs, increased use of antiplatelet drugs such as aspirin for cardiovascular disease, and side effects of antidepressants such as selective serotonin reuptake inhibitors.^30^

This study had certain limitations. First, since the JMDC database mainly includes individuals aged <65 years (up to 74 years), the generalizability of our results to older individuals remains uncertain. We speculate that the absolute risk of each outcome would be higher in older BC survivors, while the relative risks (HRs) between those with and without BC could be lower in the older population, as non-BC women also experience various age-related diseases. Second, surveillance or ascertainment bias (i.e., patients with BC have more contact with the healthcare system and are therefore more likely to be identified as having certain diseases than those without BC) could partly explain the high HRs. However, most of the studied outcomes cause symptoms such as pain, which negatively affects quality of life or is life-threatening. Therefore, we expect that both insured women with and without BC would visit hospitals or clinics similarly if they had the condition, unlike asymptomatic diseases identified through examinations (e.g., liver and kidney diseases). Third, information regarding the BC stage was unavailable, which may have influenced some outcomes, such as anxiety/depression. Instead of the BC stage, we conducted additional analyses based on treatment regimens (chemotherapy and hormone therapy), which are expected to correlate with the BC stage and can serve as proxies. Finally, there may be unmeasured or residual confounding factors, such as deprivation, diet, and exercise.

## Conclusions

In this large claims database, female BC survivors in Japan had increased risks of heart failure, atrial fibrillation/atrial flutter, and non-cardiovascular diseases (major osteoporotic fractures, other fractures, gastrointestinal bleeding, infectious pneumonia, urinary tract infection, and anxiety/depression) compared with the general population of women of the same age. We found that the risk of most diseases increased during the first year after the BC diagnosis, while the risk of fractures increased later during the follow-up. It is important for healthcare providers and patients to understand the risks of these diseases and link them to screening, prevention, and early treatment.

## Data Availability

We obtained data from the JMDC Co., and we are not allowed to share these data with other parties. Researchers who meet the criteria for access can acquire de-identified participant data from the JMDC Co. (https://www.jmdc.co.jp/en/).

## Authors’ contributions

MI and CK designed the study, conducted data processing, analysed the data, and wrote the initial draft. MI was also responsible for the funding acquisition, project administration, and supervision. KB, TK, YS, HB, TS, SN, MA, AW, and NT substantially contributed to study design, interpretation of the data, and revision of the manuscript. All authors approved the final version and had final responsibility for the decision to submit for publication.

## Acknowledgments

We thank Editage (www.editage.com) for the English language editing.

## Research in context

### Evidence before this study

The risks of non-cancer diseases among female breast cancer survivors have been investigated in the last decade. We searched PubMed on October 1, 2024, using the search terms (“cancer survivor” OR “breast cancer”) AND (“cardiovascular” OR “myocardial infarction” OR “heart failure” OR “arrhythmia” OR “atrial fibrillation” OR “stroke” OR “intracranial hemorrhage” OR “pulmonary embolism” OR “fracture” OR “gastrointestinal bleeding” OR “bleeding” OR “pneumonia” OR “urinary tract infection” OR “infection” OR “mental” OR “anxiety” OR “depression”), without restricting language or the start and end dates of the search, and conducted manual searches of reference lists from previous studies. We identified 34 studies that compared individuals with breast cancer to matched individuals without breast cancer (**Appendix pp 2–10**). We found that most studies on cardiovascular diseases were conducted in European or American countries and that there is a lack of data on other common diseases, such as bleeding and infections (**Appendix p 11**).

### Added value of this study

In the present matched-cohort study in Japan, breast cancer survivors showed an increased risk for heart failure and atrial fibrillation/flutter among cardiovascular diseases, as well as major osteoporotic fractures, other fractures, gastrointestinal bleeding, infectious pneumonia, urinary tract infections, and anxiety/depression among non-cardiovascular diseases. This study also found that the risk of most outcomes is more likely to increase during the first year, while the risk of fractures tends to rise later. To the best of our knowledge, this is the first large-scale study from Asia that investigated the risk of individual cardiovascular diseases in breast cancer survivors compared with the general population. Moreover, this is the first study to show that breast cancer survivors have a higher risk of developing bleeding and infections.

### Implications of all the available evidence

Breast cancer survivors, regardless of race or ethnicity, are at risk for a variety of non-cancer diseases, including both cardiovascular and non-cardiovascular diseases. For breast cancer survivors to live longer with a maintained quality of life, it is important for healthcare providers and patients to understand the risks of non-cancer diseases according to follow-up time (e.g., heart failure and anxiety/depression during the first year and fractures later) and to link them to screening (e.g., regular assessment of blood cholesterol, cardiac function, and bone density), prevention, and early treatment (e.g., statins, bisphosphonates, and counseling for anxiety/depression).

