## Appendix for "Cardiovascular and non-cardiovascular risks among female breast cancer survivors in Japan: A matched cohort study"

Supplementary Table S1. Summary of previous studies

Supplementary Table S2. Distribution of previous studies for each outcome by region

Supplementary Table S3. Code lists to define each outcome and covariate

Supplementary Table S4. Treatment details of the breast cancer survivors during the first year

Supplementary Figure S1. Cumulative incidence curves plotted on a logarithmic scale

Supplementary Table S5. Results of sensitivity analysis starting the follow-up from the month of breast cancer surgery

Supplementary Table S6. Incidence rate of each outcome in each group and its difference and ratio between the groups with and without breast cancer during <1 year from the index month

Supplementary Table S7. Incidence rate of each outcome in each group and its difference and ratio between the groups with and without breast cancer during 1–10 years from the index month

Supplementary Figure S2. Additional analysis by treatment regimens

**Supplementary Table S1. Summary of previous studies**

| Ref. No. | Study (author, year) | Country | Age profile | Study size | Follow-up time | Outcome and results |
|---|---|---|---|---|---|---|
| 1 | Armenian, 2016 | USA | Median 60y (range 40–96) | 36,232 cancer survivors (10,429 Breast cancer survivors including stage IV patients), 73,545 matched control (age, sex, and region-matched cancer-free controls) | 4.4y exposed, 4.5y matched cohort | Ischemic heart disease; crude IRR 1.09 (0.97–1.22), aIRR 1.04 (0.93–1.16),<br>Cardiomyopathy/heart failure; crude IRR 1.51 (1.33–1.71), aIRR 1.35 (1.19–1.54),<br>Stroke; IRR 1.12 (1.00–1.26), aIRR 1.07 (0.95–1.20) |
| 2 | Abdel-Qadir, 2019 | Canada | Median 61y | 78,318 breast cancer survivors (stage I–III), 2,394,954 controls (age-matched cancer-free controls) | 5.7y exposed | Ischemic heart disease (hospitalization); HR 1.12 (1.07–1.19), aHR 0.99 (0.94–1.05),<br>Heart failure (hospitalization); HR 1.41 (1.33–1.50), aHR 1.21 (1.14–1.29),<br>Cerebrovascular disease (hospitalization); HR 1.23 (1.16–1.30), aHR 1.10 (1.04–1.17)<br>Arrhythmia (hospitalization); HR 1.49 (1.40–1.59), aHR 1.31 (1.23–1.39) |
| 3 | Jordan, 2014 | USA | restricted to age ≥ 65y | 1,361 breast cancer survivors (stage I–II, 5 years survivors), 1,361 controls (age and health system-matched cancer-free controls) | 3.3y exposed, 3.7y controls | Myocardial infarction; aHR 0.75 (0.57–0.99)<br>Congestive heart failure; aHR 0.92 (0.76–1.1)<br>Cerebrovascular disease ; aHR 0.85 (0.68–1.1)<br>Pulmonary embolism ; aHR 0.61 (0.29–1.3)<br>Osteoporotic fracture: aHR 1.0 (0.83–1.2) |

| | | | | | | |
|---|---|---|---|---|---|---|
| 4 | Rugbjerg, 2016 | Denmark | restricted to age 15-39y, median age group 30-34y | 43,153 cancer survivors (5,731 Breast cancer survivors defined as ICD-10 C50, 1-year survivor), 255,513 controls (sex and age-matched) | exposed mean 15 y (range 0–34 y) | Myocardial infarction (hospitalization); RR 1.12 (0.92–1.36),<br>Heart failure (hospitalization); RR 1.53 (1.30–1.80),<br>Cerebrovascular disease; RR 1.16 (1.01–1.34),<br>Arrhythmia (hospitalization); RR 1.26 (1.09–1.45),<br>Pulmonary embolism (hospitalization); RR 1.83 (1.36–2.48) |
| 5 | Matthews, 2021 | USA | restricted to age ≥ 65y | 91, 473 breast cancer survivors (stage I–III), 454,197 controls (age, SEER region, and race/ethnicity matched cancer-free controls) | Not reported | Myocardial infarction; Rate difference -0.95 (-2.00 – 0.10), HR 0.98 (0.95–1.00), aHR 0.97 (0.94–1.00)<br>Heart failure; Rate difference 14.49 (11.74–17.24), HR 1.08 (1.07–1.10), aHR 1.07 (1.06–1.09)<br>Stroke; Rate difference 0.75 (0.99–0.98), HR 0.99 (0.98–1.01), aHR 0.98 (0.97–1.00)<br>Arrhythmia ; Rate difference 0.75 (0.99–0.98), HR 0.99 (0.98–1.01), aHR 1.07 (1.06–1.09)<br>Pulmonary embolism; Rate difference 0.12 (-0.16 – 0.40), HR 1.03(0.93–1.15), aHR 0.98 (0.87–1.10) |
| 6 | Kero, 2014 | Finland | restricted to age ≤35y | 9,401 cancer survivors (survived at least 5y, 919 breast cancer), 43,392 siblings (without early-onset cancer) | Not reported | Myocardial infarction/cardiac ischemia; HR0.9 (0.6–1.5),<br>Cardiac arrhythmia; HR1.1 (0.7–1.8) |
| 7 | Boerman, 2014 | Netherland | restricted to age ≤80y, Median 56y (Radiotherapy group), 47y (Chemotherapy | 561 breast cancer survivors (all received surgery), 1,635 controls (age-matched cancer-free controls) | median 9y (range 5–57 years) | Congestive heart failure; HR 0.5 (0.2–1.8) for Radiotherapy patients, HR 1.8 (0.6–5.8) for chemotherapy patients |

| | | | | | | |
|---|---|---|---|---|---|---|
| | | | group), 54y (controls) | | | |
| 8 | Brand, 2017 | Sweden | Restricted to ages 25-75, mean 57y | 8,338 breast cancer survivors, 39,013 controls (age-matched) | exposed 7.2y, controls 5.9y | Pulmonary embolism; HR 3.09 (2.50–3.81) |
| 9 | Strongman, 2019 | UK | patients aged ≥18 years | 126,120 cancer survivors (25,633 breast cancer survivors defined as ICD-10 C50), 630144 controls (matched by age (±3 years), sex, and general practice) | exposed 5.7y, controls 6.4y | Myocardial infarction; HR 0.85 (0.76–0.96), aHR 0.82 (0.73–0.93)<br>Heart failure; HR 1.14 (1.06–1.23), aHR 1.13 (1.04–1.23),<br>Arrhythmia; HR 1.04 (0.98–1.10), aHR 1.04 (0.98–1.10)<br>Stroke; HR 1.08 (1.00–1.17), aHR 1.07 (0.99–1.16)<br>Pulmonary embolism; HR 2.26 (2.02–2.53), aHR 2.29 (2.04–2.57) |
| 10 | Yeh, 2022 | Taiwan | restricted to age ≥ 20y, mean 60.6y | 552,485 cancer survivors (111,273 breast cancer survivors defined as ICD-10 C50), controls (matched by age and sex) | exposed 4.1y (range 1.7–7.6) | Cardiovascular disease; HR 1.14 (1.09–1.19), aHR (age and sex) 1.15 (1.09–1.20) |
| 11 | Raisei-Estabragh, 2023 | UK | mean 62y | 18,714 cancer survivors (9,531 breast cancer survivors defined as ICD-10 C50), 18,714 controls (propensity score matching using age, sex, ethnicity, Townsend score, education, blood pressure, heart rate, BMI, HbA1c, glucose, cholesterol, HDL, LDL, Triglyceride, | 11.8±1.7 years | Ischemic heart disease; aHR 1.05 (0.93–1.19)<br>Heart failure; aHR 1.34 (1.14–1.57)<br>Atrial fibrillation/flutter; aHR 1.11 (0.98–1.25)<br>Stroke; aHR 1.13 (0.91–1.38) |

| | | | | | | |
|---|---|---|---|---|---|---|
| | | | | physical activity, smoking status, diabetes, hypertension, high cholesterol) | | |
| 12 | Koric, 2022 | USA | ≥18 y of age | 6,641 breast cancer survivors (survived at least 10 years), 36,612 matched controls (matched by age and state) | Not reported | Acute myocardial infarction; 10-15y HR 1.02 (0.62–1.68), >15y HR 0.20 (0.02–2.78)<br>Heart failure; 10-15y HR 0.96 (0.71–1.31), >15y HR 0.54 (0.16–1.80)<br>Stroke; 10-15y HR1.24 (1.00–1.53), >15y HR 0.90 (0.54–1.35) |
| 13 | Navi, 2015 | USA | ≥ 66 y | 327,389 pairs of cases and controls (61,118 breast cancer survivors, controls were matched by age, sex, race, SEER registry, and Charlson comorbidity index in the year prior to study entry) | Not reported | Stroke; 1-3M aHR 1.17 (1.03–1.32), 9–12 M aHR 0.93 (0.83–1.04) |
| 14 | Yang, 2022 | Sweden | age 25–75 y | 8015 breast cancer patients, 80,072controls (matched by age) | 10.8 years | Ischemic heart disease; <1y 1.45 (1.03–2.04), >10y HR 0.79 (0.61–1.03)<br>Heart failure; <1y HR 2.71 (1.70–4.33), >10y HR 1.28 (1.03–1.59)<br>Arrhythmia; <1y HR 2.14 (1.63–2.81), >10y HR 1.42 (1.21–1.67) |
| 15 | Greenlee, 2022 | US | age ≥ 21 y, mean 60.3y | 13642 breast cancer survivors (stage I-IV and had a history of chemotherapy, radiation therapy, or endocrine therapy), 68,202 controls (matched by age and | Over a 7-year average follow-up (range < 1-14 years) | Heart failure/cardiomyopathy<br>Anthracyclines without Transtuzumab; aHR 1.84 (1.21–2.80),<br>Anthracyclines with Transtuzumab; aHR 3.68 (1.79–7.59),<br>Transtuzumab without anthracyclines; aHR 2.53 (1.33– |

| | | | | | | |
|---|---|---|---|---|---|---|
| | | | | race/ethnicity) | | 4.81),<br>Radiation therapy; aHR 1.38 (1.13–1.69),<br>Aromatase Inhibitor ; aHR 1.31 (1.07–1.60),<br>Tamoxifen ; aHR 1.39 (0.82–2.35) |
| 16 | Tsai, 2013 | Taiwan | Breast cancer survivors mean 51.2y, Controls mean 51.4y | 22,076 breast cancer patients, 88,304 comparison women without cancer (matched by age and index date) | Not reported | All fractures (non-traumatic); aHR 1.29 (1.11–1.51),<br>Hip fractures (non-traumatic); aHR 1.37 (1.08–1.74),<br>Distal forearm (non-traumatic); aHR 1.09 (0.84–1.43),<br>Vertebral (non-traumatic); aHR 1.48 (1.07–2.04) |
| 17 | Pawloski, 2013 | US | age ≥ 65y, mean age 77.7y | 1,286 breast cancer survivors (survived at least 5 years), 1,286 comparison women (matched by age, study site, and enrollment year) | survivors 6.0 y, comparison 6.5 y | All fractures (hip, wrist, and vertebrae); aHR1.1 (0.9–1.3),<br>Hip and wrist fractures (hip and wrist); aHR1.1 (0.9–1.4) |
| 18 | Rees-Punia, 2022 | US | mean 69.4y | 14,159 cancer survivors (numbers of breast cancer survivors are unknown), 78272 controls | Not reported | Pelvic fracture; 1-5y aHR 0.78 (0.51–1.18), >5y aHR 0.96 (0.64–1.44),<br>Vertebral fracture; 1-5y aHR 0.65 (0.46–0.91), >5y aHR 0.89 (0.62–1.28)<br>Radial fracture; 1-5y aHR 0.61 (0.38–0.97), >5y aHR 1.07 (0.60–1.91) |
| 19 | Gong, 2023 | Canada | 29.4% of the population was aged ≥ 65 y in both groups | 172,963 cancer survivors (56,424 breast cancer survivors defined as ICD-10 C50), 172,963 non-cancer controls (matched by age and sex) | 6.5y | Fracture ; age<65y aHR 1.44 (1.35–1.53), age≥ 65y aHR 1.05 (0.96–1.16)<br>Spine and pelvis fracture; aHR 1.41 (1.18–1.67),<br>Hip and femur fracture; aHR 1.21 (0.99–1.31),<br>Leg fracture; aHR 1.23 (1.11–1.37),<br>Wrist and shoulder fracture; aHR 1.25 (1.16–1.34) |

| | | | | | | |
|---|---|---|---|---|---|---|
| 20 | Go, 2020 | Korea | Breast cancer survivors 54.0±5.6y, Controls 54.3±6.7y | 74 breast cancer survivors (stage I–III, survived at least 5 years), 296 non-cancer controls (propensity score matching by age and survey year) | Not reported | fracture incidence of breast cancer survivors was 2.7%, non-cancer controls had a fracture incidence of 9.2% (P = 0.146). |
| 21 | Stumpf, 2019 | UK | aged between 18 and 50, mean 43.3y | 1,761 breast cancer survivors (defined as ICD-10 C50), 1,761 controls (matched by age, index year, and physician) | Not reported | Fracture incidence<br>Breast cancer vs. no cancer aHR 2.39 (1.70–3.34),<br>Breast cancer with tamoxifen vs. no cancer aHR 2.67 (1.58–4.53),<br>Breast cancer without endocrine therapy vs. no cancer aHR 1.63 (0.80–3.33) |
| 22 | Stumpf, 2020 | UK | mean 68.6 y | 4,115 breast cancer survivors (defined as ICD-10 C50), 4,115 controls (matched by age, index year, and physician) | Not reported | Fracture incidence<br>Breast cancer vs. matched healthy cohort aHR 2.44 (1.99–2.98),<br>Breast cancer and tamoxifen vs. matched healthy cohort aHR 0.63 (0.34–1.17),<br>Breast cancer and aromatase inhibitors vs. matched healthy cohort aHR 3.36 (2.65–4.26),<br>Breast cancer and chemotherapy vs. matched healthy cohort aHR 0.88 (0.60–1.28) |

| | | | | | | |
|---|---|---|---|---|---|---|
| 23 | Yang, 2022 | Taiwan | breast cancer survivors mean 52.3 y, controls mean 51.7 y | 96,862 breast cancer survivors, 96,862 non-cancer controls (propensity score matching using age, income, urbanization level of residential areas, diagnosis year, and Charlson comorbidity index) | Not reported | Anxiety; aHR 1.29 (1.25–1.33),<br>Depression; aHR 1.78 (1.69–1.87) |
| 24 | Khan, 2010 | UK | age ≥ 30y | 16,938 breast cancer survivors, 67,649 controls without cancer | Not reported | Depression; aOR 1.06 (1.00–1.12)<br>Anxiety; aOR 1.06 (0.97–1.16) |
| 25 | Hung, 2013 | Taiwan | age ≥ 20y,<br>Median 49y | 26,629 breast cancer survivors, 26629 controls (matched by age, sex, and presence of comorbidities with the same diagnosis index date) | breast cancer survivors 2.70y, the matched group 3.22 y | Depression; RR 1.94 (1.76–2.18)<br>Anxiety; RR 1.22 (1.16–1.29) |
| 26 | Earie, 2007 | USA | breast cancer survivors mean 60.2 y, controls mean 60.5 y | 1,111 cancer survivors (468 breast cancer survivors), 4444 controls (matched by age, sex, and clinic location.) | breast cancer survivors 2.5y, the matched group 1.7 y | Diagnosis of Anxiety/sleep disorder was 19.0% in breast cancer survivors, 17.2% for Female controls |
| 27 | Kim, 2017 | Korea | age ≥ 16 y | 2,130 breast cancer survivors who have undergone mastectomy, 8,520 controls (matched by age, sex, income, region, and pre-operative depression) | Not reported | Diagnosis of Anxiety/sleep disorder;<br>For <1y, 4.8% in breast cancer survivors, 3.1% for Female controls (p<0.001)<br>For after <4y, no difference between the two groups. |

| | | | | | | |
|---|---|---|---|---|---|---|
| 28 | Carreira, 2021 | UK | age ≥ 18 y | 57,571 breast cancer survivors, 230,067 non-cancer women (age, primary care practice, and eligibility of the data for linkage to hospital data sources matched) | Exposed4.5y, comparison group 5.2y | Depression; aHR 1.35(1.32–1.38)<br>Anxiety; aHR 1.35(1.32–1.38) |
| 29 | Tran, 2020 | Korea | breast cancer survivors mean 57.3 y, controls mean 57.2 y | 273 breast cancer survivors (stage 0–II who had surgery, mean survival time 10 y), 819 controls (matched by age, education-matched) | Not reported | Pain/discomfort (46% vs. 23%, p<0.001),<br>Anxious/depressed feelings (44% vs. 8%, p<0.001) |
| 30 | Buzasi, 2023 | UK | age ≥ 18 y (breast cancer survivors mean 61.7y, controls mean 61.8y) | 578,160 cancer survivors (145,515 breast cancer survivors), 3,226,404 controls (matched by index date, year of birth (±3 years), sex, and primary care practice) | Breast cancer group mean 6.65y, controls mean 7.35y | Any bone fracture; 1 to <2y 1.22 (1.16–1.29), 2 to <5y 1.25 (1.20–1.29), ≥5y 1.17 (1.14–1.21)<br>Major bone fracture; 1 to <2y 1.31 (1.21–1.43), 2 to <5y 1.26 (1.19–1.33), ≥5y 1.23(1.17–1.28) |
| 31 | Guha, 2022 | USA | age ≥ 66 y | 85,423 breast cancer patients, controls (1:1 matched by year of birth, race, SEER registry, and Charlson comorbidity index | Not reported | 1y atrial fibrillation incidence of breast cancer survivors was 3.3%, 1y atrial fibrillation incidence of control was 1.8% |
| 32 | D’Souza, 2019 | Denmark | breast cancer survivors median 62 y, controls median 62 y | 74,155 breast cancer survivors, 222,465 controls (matched by age and sex) | Median 3 y | Atrial fibrillation (<60y); <6 month HR 2.10 (1.25–3.44), 6 to 3y HR1.80 (1.38–2.35)<br>Atrial fibrillation (>60y); <6 month HR 1.13 (0.95–1.34), 6 to 3y HR1.14 (1.05–1.25) |

| | | | | | | |
|---|---|---|---|---|---|---|
| 33 | Yun, 2021 | Korea | cancer survivors mean 57.5 y, controls mean 57.5 y | 816,811 cancer survivors (80,920 breast cancer survivors), 1,633,663 controls (matched by age and sex) | Median 4.5 y | Atrial fibrillation; <90 days aHR 1.48 (1.39–1.58), <1y aHR 1.40 (1.30–1.50), <5y aHR 1.00 (0.84–1.18) |
| 34 | Park, 2024 | Korea | age ≥ 18 y, cancer survivors mean 51.6 y, controls mean 51.6 y | 113,232 breast cancer survivors (who underwent surgery), 566,160 controls (matched by age) | Breast cancer group mean 5.1y, control group 5.3y | Atrial fibrillation; all age aHR 1.06 (1.00–1.13) 18–39y aHR 2.79 (1.98–3.94), 40-50y aHR 1.22 (1.03–1.45), 51–65y aHR 1.13 (1.03–1.25), ≥ 66y 0.90 (0.81–0.99) |

IRR = incidence rate ratio, HR = hazard ratio, RR = rate ratio, aHR = adjusted hazard ratio, aOR = adjusted odds ratio.

**Supplementary Table S2. Distribution of previous studies for each outcome by region**

<table>
<tr><th>Outcome</th><th>USA</th><th>Europe</th><th>Asia</th><th>Other regions</th></tr>
<tr><td>Myocardial infarction</td><td>1, 3, 5, 12</td><td>4 (Denmark), 6 (Finland), 9 (UK), 11 (UK), 14 (Sweden)</td><td rowspan="5">10 (Taiwan): cardiovascular disease,<br>33 (Korea): atrial fibrillation,<br>34 (Korea): atrial fibrillation</td><td>2 (Canada)</td></tr>
<tr><td>Heart failure</td><td>1, 3, 5, 12, 15</td><td>4 (Denmark), 7 (Netherland), 9 (UK), 11 (UK), 14 (Sweden),</td><td>2 (Canada)</td></tr>
<tr><td>Atrial fibrillation/flutter</td><td>5, 31</td><td>4 (Denmark), 6 (Finland), 9 (UK), 11 (UK), 14 (Sweden), 32 (Denmark)</td><td>2 (Canada)</td></tr>
<tr><td>Stroke</td><td>1, 3, 5, 12, 13</td><td>4 (Denmark), 9 (UK), 11 (UK)</td><td>2 (Canada)</td></tr>
<tr><td>Pulmonary embolism</td><td>3, 5</td><td>4 (Denmark), 8 (Sweden), 9 (UK)</td><td>None</td></tr>
<tr><td>Fractures</td><td>3, 17, 18</td><td>21 (UK), 22 (UK), 30(UK)</td><td>16 (Taiwan), 20 (Korea)</td><td>19 (Canada)</td></tr>
<tr><td>Gastrointestinal bleeding</td><td>None</td><td>None</td><td>None</td><td>None</td></tr>
<tr><td>Infectious pneumonia</td><td>None</td><td>None</td><td>None</td><td>None</td></tr>
<tr><td>Urinary tract infection</td><td>None</td><td>None</td><td>None</td><td>None</td></tr>
<tr><td>Anxiety/depression</td><td>26</td><td>24 (UK), 28 (UK)</td><td>23(Taiwan), 25 (Taiwan), 27 (Korea), 29 (Korea)</td><td>None</td></tr>
</table>

USA = United States of America, UK = United Kingdom.

Note: Each number is corresponding to the reference number in Supplementary Table S1.

**Supplementary Table S3. Code lists to define each outcome and covariate**

| Outcomes/Covariates | Diagnosis (International Classification of Diseases and Related Health Problems, 10th Revision code) | Medication (ATC code or EphMRA code) | Medical procedures (Japanese original code) |
|---|---|---|---|
| Myocardial infarction | I21 (I21.0, I21.01, I21.2, I21.3, I21.4, I21.9), I24.9 | | K546, K548, K549, K552, K552–2 |
| Heart failure | I50 (I50.0, I50.1, I50.9), I11.0, I13.0, I13.2 | ATC code; C01CA04, C01CA07<br>EphMRA code; C03 | “Echocardiogram” |
| Atrial fibrillation/Atrial flutter | I48 (I48.0, I48.1, I48.2, I48.3, I48.4, I48.9) | ATC code; B01AA03, B01AE07, B01AF01, B01AF02, B01AF03<br>EphMRA code; C01B | K595, K594 |
| Ischemic stroke | I63 (I63.0, I63.1, I63.2, I63.3, I63.4, I63.5, I63.6, I63.8, I63.9) | ATC code; B01AA03, B01AC04, B01AC06, B01AC23, B01AD02, B01AE03, B01AE07, B01AF01, B01AF02, B01AF03, N07XX14<br>EphMRA code; B01B1, B01B2 | K149, K164-3, K178-3, K178-4 |
| Intracranial hemorrhage | I60 (I60.0, I60.1, I60.2, I60.3, I60.4, I60.5, I60.6, I60.7, I60.8, I60.9), I61 (I61.0, I61.1, I61.3, I61.4, I61.5, I61.6, I61.9), I62 (I62.0, I62.1, I62.9) | ATC code; C02DD01, C01DA02, C08CA04, C08DB01 | K164, K164-4, K164-5, K177, K178 |
| Pulmonary embolism | I26 (I26.0, I26.9) | ATC code; B01AA03, B01AD02, B01AE07, B01AF01, B01AF02, B01AF03<br>EphMRA code; B01B1, B01B2 | K592, K620 |
| Major osteoporotic fractures | M80 (M80.0, M80.1, M80.2, M80.3, M80.4, M80.5, M80.8, M80.9), S22.0, S22.1, S32 (S32.0, S32.1, S32.2, S32.3, S32.4, S32.5, S32.7, S32.8), S42.2, S52.5, S52.6, S72.0, T02.1 | | "Surgery of the musculoskeletal system, extremities, and trunk", "Orthopedic procedures (e.g., traction, Chest fixation band)", "Orthopedic cast" |

| | | | |
|---|---|---|---|
| Other fractures | S02 (S02.0, S02.1, S02.2, S02.3, S02.4, S02.5, S02.6, S02.7, S02.8, S02.9), S12 (S12.0, S12.1, S12.2, S12.7, S12.8, S12.9), S22.2, S22.3, S22.4, S22.5, S22.8, S22.9, S42.0, S42.1, S42.3, S42.4, S42.7, S42.8, S42.9, S52.0, S52.1, S52.2, S52.3, S52.4, S52.7, S52.8, S52.9, S62 (S62.0, S62.1, S62.2, S62.3, S62.4, S62.5, S62.6, S62.7, S62.8), S72.1, S72.2, S72.3, S72.4, S72.7, S72.9, S82 (S82.0, S82.1, S82.2, S82.3, S82.4, S82.5, S82.6, S82.7, S82.8, S82.9), S92 (S92.0, S92.1, S92.2, S92.3, S92.4, S92.5, S92.7, S92.9), T02.0, T02.2, T02.3, T02.4, T02.5, T02.6, T02.7, T02.8, T02.9, T08, T10, T12, T14.2 | | "Surgery of the musculoskeletal system, extremities, and trunk", "Orthopedic procedures (e.g., traction, Chest fixation band)", "Orthopedic cast" |
| Gastrointestinal bleeding | I85.0, K22.6, K25.0, K25.1, K25.2, K25.4, K25.5, K25.6, K26.0, K26.1, K26.2, K26.4, K26.5, K26.6, K27.0, K27.1, K27.2, K27.4, K27.5, K27.6, K28.0, K28.1, K28.2, K28.4, K28.5, K28.6, K29.0, K92.0, K92.1, K92.2 | | D306, D308, D310, D312, D313 |
| Infectious pneumonia | B01.2, B05.2, B20.6, B25.0, J10.0, J11.0, J12 (J12.0, J12.1, J12.2, J12.3, J12.8, J12.9), J13, J14, J15 (J15.0, J15.1, J15.2, J15.3, J15.4, J15.5, J15.6, J15.7, J15.8, J15.9), J16 (J16.0, J16.8), J17 (J17.0, J17.1, J17.2, J17.3, J17.8), J18 (J18.0, J18.1, J18.2, J18.8, J18.9), J85.1, U04.9 | EphMRA code; J01, J02, J04, J05 | |
| Urinary tract infection | N10, N12, N13.6, N15.1, N15.9, N30.0, N30.8, N30.9, N39.0 | EphMRA code; J01, J02 | |

| | | |
|---|---|---|
| Anxiety/depression | F32 (F32.0, F32.1, F32.2, F32.3, F32.8, F32.9), F33 (F33.0, F33.1, F33.2, F33.3, F33.4, F33.8, F33.9), F34 (F34.0, F34.1, F34.8, F34.9), F40 (F40.0, F40.1, F40.2, F40.8, F40.9), F41 (F41.0, F41.1, F41.2, F41.3, F41.8, F41.9), F42 (F42.0, F42.1, F42.2, F42.8, F42.9), F43 (F43.0, F43.1, F43.2, F43.8, F43.9), F48 (F48.0, F48.1, F48.8, F48.9) | EphMRA code; N05B, N06A |
| Diabetes | E10 (E10.0, E10.1, E10.2, E10.3, E10.4, E10.5, E10.6, E10.7, E10.8, E10.9), E11 (E11.0, E11.1, E11.2, E11.3, E11.4, E11.5, E11.6, E11.7, E11.8, E11.9), E12 (E12.0, E12.1, E12.2, E12.3, E12.4, E12.5, E12.6, E12.7, E12.8, E12.9), E13 (E13.0, E13.1, E13.2, E13.3, E13.4, E13.5, E13.6, E13.7, E13.8, E13.9), E14 (E14.0, E14.1, E14.2, E14.3, E14.4, E14.5, E14.6, E14.7, E14.8, E14.9) | EphMRA code; A10 |
| Dyslipidemia | E78 (E78.0, E78.1, E78.2, E78.3, E78.4, E78.5, E78.6, E78.8, E78.9) | EphMRA code; C10, C11 |
| Hypertension | I10, I11 (I11.0, I11.9), I12 (I12.0, I12.9), I13 (I13.0, I13.1, I13.2, I13.9), I15 | EphMRA code; C03, C07, C08, C09, C11 |
| Osteoporosis | M80 (M80.0, M80.1, M80.2, M80.3, M80.4, M80.5, M80.8, M80.9), M81 (M81.0, M81.1, M81.2, M81.3, M81.4, M81.5, M81.6, M81.8, M81.9) | ATC code; M05BX08, A14AB01, S01XA11 EphMRA code; A12A-, G03C-, A11C2, M05B3, G03J-, H04A-, H04E-, M05B9, |

ATC = Anatomical Therapeutic Chemical, EphMRA = European Pharmaceutical Market Research Association.

**Supplementary Table S4. Treatment details of the breast cancer survivors during the first year**

| | No. of breast cancer survivors N=24,017 |
|---|---|
| | n (%) |
| Surgery | 24,017 (100) |
| Axillary lymph node dissection | 6,697 (27.9) |
| Radiotherapy | 12,567 (52.3) |
| Chemotherapy/molecularly targeted drug | |
| Anthracycline (doxorubicin or epirubicin) | 6,494 (27.0) |
| Taxane (docetaxel or paclitaxel) | 7,884 (32.8) |
| Anthracycline and taxane | 5,732 (23.9) |
| HER2 targeted therapy (trastuzumab or pertuzumab) | 2,860 (11.9) |
| Taxane and HER2 targeted therapy | 2,528 (10.5) |
| Hormone therapy | |
| Tamoxifen | 10,607 (44.2) |
| Aromatase inhibitor (anastrozole, letrozole, or exemestane) | 5,891 (24.5) |

HER2 = human epidermal growth factor receptor 2.

**Supplementary Figure S1. Cumulative incidence curves plotted on a logarithmic scale**

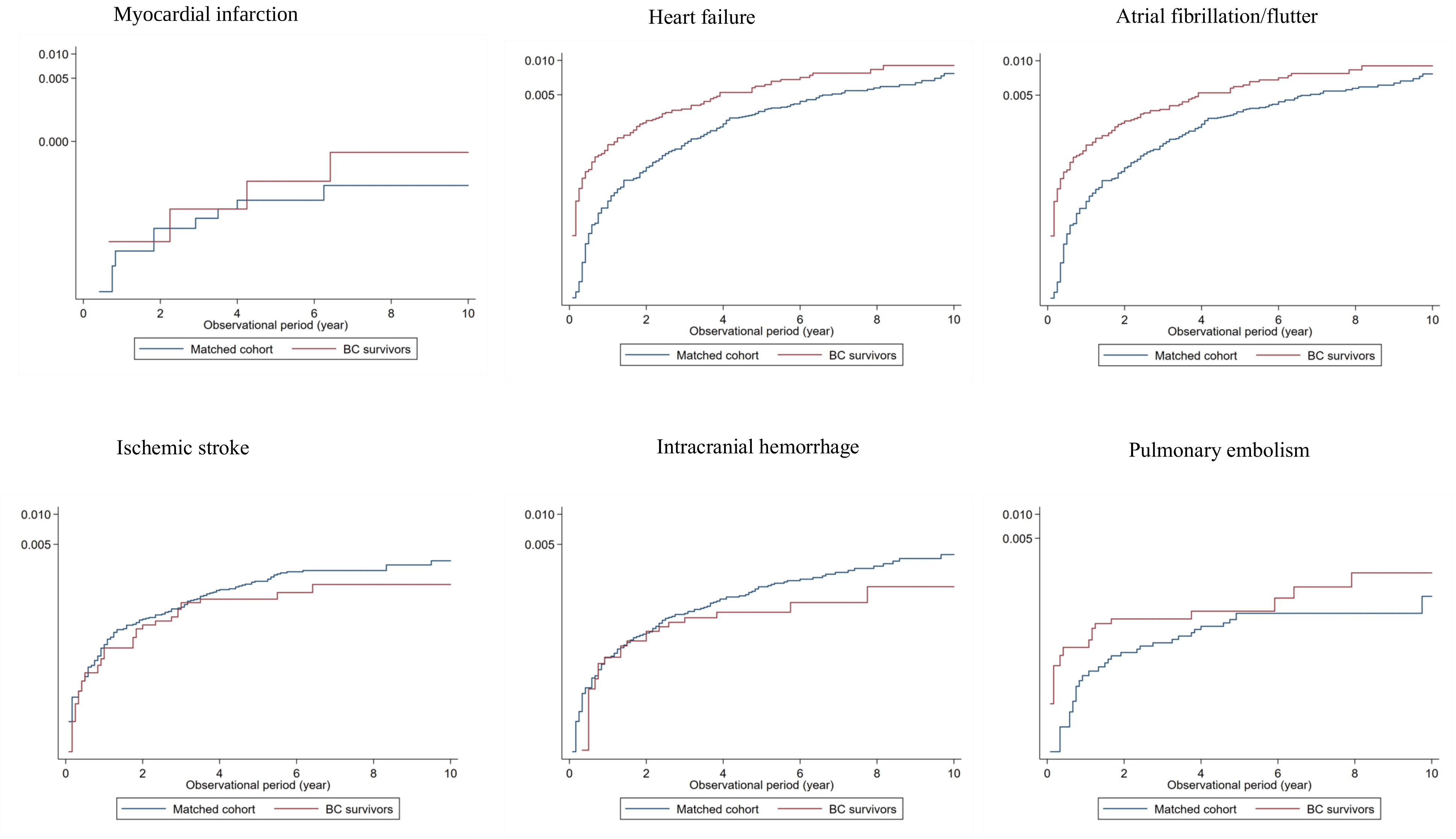

Major osteoporotic fractures

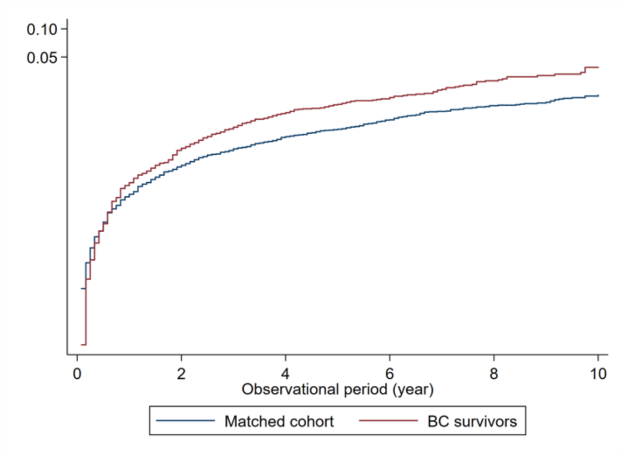

Non-osteoporotic fractures

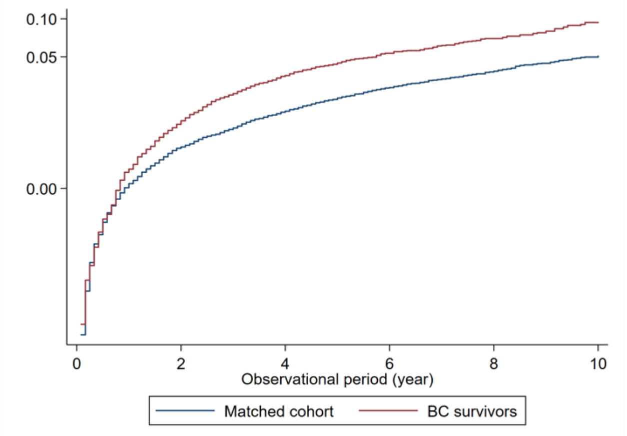

Gastrointestinal bleeding

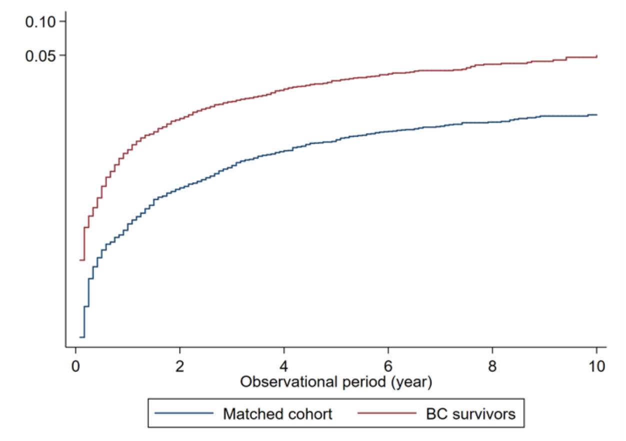

Infectious pneumonia

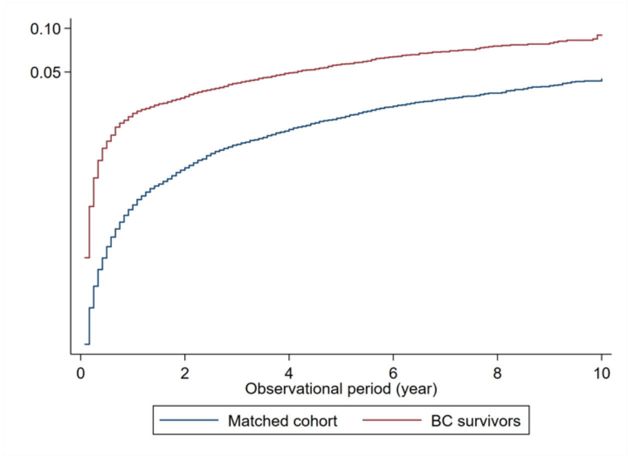

Urinary tract infection

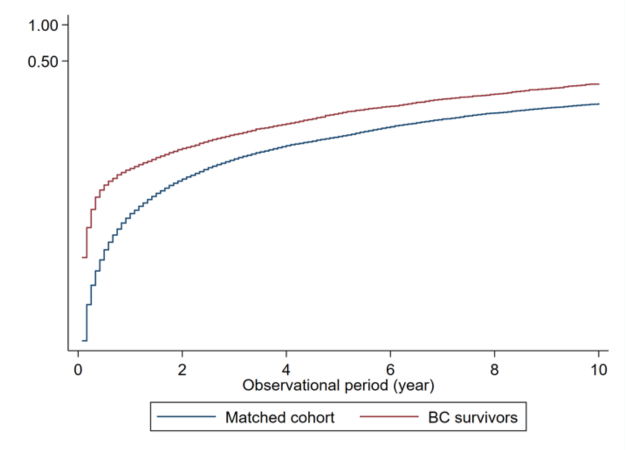

Anxiety/depression

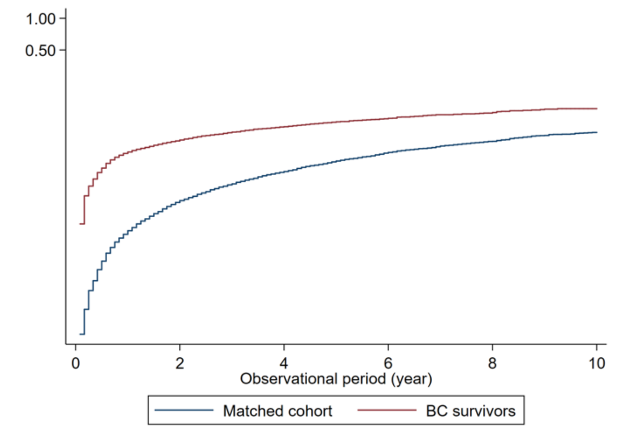

**Supplementary Table S5. Results of sensitivity analysis starting the follow-up from the month of breast cancer surgery**

| | Patients with breast cancer | | Women without breast cancer | | Rate difference (95% CI) (/1000 PY) | Rate ratio (95% CI) | Hazard ratio (95% CI) | | | |
|---|---|---|---|---|---|---|---|---|---|---|
| | No. of outcomes/ No. of patients* | Rate (95% CI) (/1000 PY) | No. of outcomes/ No. of patients* | Rate (95% CI) (/1000 PY) | | | Model 1 | Model 2 | Model 3 | Model 4 |
| Myocardial infarction | 4/23,659 | 0.05 (0.02–0.15) | 9/93,300 | 0.03 (0.02–0.06) | 0 (-0.03–0.02) | 0.87 (0.33–2.19) | 1.35 (0.40–4.58) | NA | NA | NA |
| Heart failure | 50/23,692 | 0.68 (0.52–0.90) | 77/93,244 | 0.27 (0.21–0.33) | 0.41 (0.22–0.61) | 2.56 (1.76–3.70) | 3.16 (2.13–4.68) | 3.21 (2.10–4.91) | 3.04 (1.36–6.82) | 3.42 (2.19–5.33) |
| Atrial fibrillation/flutter | 83/23,576 | 1.13 (0.91–1.41) | 197/93,038 | 0.68 (0.59–0.78) | 0.45 (0.19–0.71) | 1.66 (1.27–2.16) | 1.68 (1.28–2.21) | 1.63 (1.23–2.15) | 1.57 (1.01–2.44) | 1.63 (1.23–2.16) |
| Ischemic stroke | 22/23,646 | 0.30 (0.20–0.45) | 113/93,180 | 0.38 (0.31–0.47) | -0.09 (-0.24–0.05) | 0.77 (0.46–1.22) | 0.73 (0.45–1.18) | 0.67 (0.41–1.11) | 0.86 (0.40–1.86) | 0.67 (0.41–1.12) |
| Intracranial hemorrhage | 19/23,651 | 0.26 (0.16–0.41) | 103/93,220 | 0.36 (0.29–0.45) | -0.09 (-0.23– -0.04) | 0.54 (0.26–1.02) | 0.73 (0.42–1.19) | 0.65 (0.38–1.11) | 0.99 (0.40–2.46) | 0.64 (0.38–1.11) |
| Pulmonary embolism | 11/23,651 | 0.15 (0.08–0.27) | 26/93,288 | 0.09 (0.06–0.13) | 0.06 (-0.04–0.15) | 1.86 (0.74–3.49) | 1.64 (0.79–3.39) | NA | NA | NA |

| | | | | | | | | | | |
|---|---|---|---|---|---|---|---|---|---|---|
| Major osteoporotic fractures | 231/23,484 | 3.18 (2.79–3.61) | 514/92,775 | 1.79 (1.64–1.94) | 1.39 (0.95–1.83) | 1.78 (1.52–2.08) | 1.68 (1.42–1.99) | 1.68 (1.42–1.99) | 1.57 (1.22–2.03) | 1.67 (1.42–1.99) |
| Other fractures | 622/23,027 | 8.82 (8.15–9.54) | 1,368/91,677 | 4.73 (4.44–5.03) | 4.74 (3.86–5.61) | 2.00 (1.80–2.23) | 1.85 (1.67–2.05) | 1.85 (1.67–2.05) | 1.89 (1.64–2.18) | 1.86 (1.68–2.06) |
| Gastrointestinal bleeding | 418/23,292 | 5.49 (4.90–6.15) | 481/92,642 | 1.68 (1.53–1.83) | 4.17 (3.59–4.75) | 3.49 (3.05–3.98) | 3.53 (3.07–4.05) | 3.51 3.05–4.04) | 3.18 (2.58–3.93) | 3.51 (3.05–4.05) |
| Infectious pneumonia | 781/22,752 | 11.4 (10.6–12.2) | 1,372/90,607 | 4.93 (4.67–5.20) | 6.43 (5.59–7.27) | 2.30 (2.11–2.52) | 2.45 (2.32–2.59) | 2.37 (2.16–2.56) | 2.33 (2.02–2.69) | 2.38 (2.16–2.61) |
| Urinary tract infection | 2,094/20,383 | 35.6 (34.1–37.1) | 5,930/82,654 | 24.2 (23.6–24.9) | 11.4 (9.71–13.0) | 1.47 (1.40–1.54) | 1.49 (1.41–1.57) | 1.48 (1.40–1.56) | 1.45 (1.34–1.57) | 1.48 (1.40–1.56) |
| Anxiety/depression | 1,183/21,424 | 18.6 (17.5–19.6) | 2,335/87,568 | 8.70 (8.35–9.06) | 9.86 (8.75–11.0) | 2.13 (1.99–2.29) | 2.23 (2.07–2.41) | 2.24 (2.08–2.42) | 2.30 (2.06–2.57) | 2.25 (2.09–2.43) |

CI = confidence interval, NA = not available (because the number of outcomes was too small).

*Patients with a previous history of the outcome were excluded from the analysis.

Model 1: without additional adjustment

Model 2: adjusted for hypertension, diabetes, and dyslipidemia, in addition to osteoporosis for the fracture outcome

Model 3: further adjusted for BMI, smoking history, and drinking habits as a complete case analysis

Model 4: with multiple imputation for BMI, smoking history, and drinking habits

**Supplementary Table S6. Incidence rate of each outcome in each group and its difference and ratio between the groups with and without breast cancer during <1 year from the index month**

| | Patients with breast cancer | | Women without breast cancer | | Rate difference (95% CI) (/1000 PY) | Rate ratio (95% CI) | Hazard ratio (95% CI) | | | |
|---|---|---|---|---|---|---|---|---|---|---|
| | No. of outcomes/ No. of patients* | Rate (95% CI) (/1000 PY) | No. of outcomes/ No. of patients* | Rate (95% CI) (/1000 PY) | | | Model 1 | Model 2 | Model 3 | Model 4 |
| Myocardial infarction | 1/24,011 | 0.05 (0-0.31) | 3/96,059 | 0.03 (0.01–0.11) | 0.01 (-0.09–0.11) | 1.32 (0.03–16.4) | 1.15 (0.12–11.1) | NA | NA | NA |
| Heart failure | 24/23,989 | 1.09 (0.73–1.62) | 23/95,999 | 0.26 (0.17–0.39) | 0.82 (0.38–1.27) | 4.12 (2.23–7.64) | 4.03 (2.26–7.21) | 4.73 (2.49–8.97) | 13.3 (2.25–78.4) | 5.59 (2.31–13.5) |
| Atrial fibrillation/flutter | 41/23,938 | 1.86 (1.37–2.53) | 51/95,791 | 0.59 (0.45–0.77) | 1.28 (0.68–1.87) | 3.18 (2.05–4.89) | 3.16 (2.08–4.80) | 3.20 (2.10–4.90) | 2.60 (1.33–5.11) | 3.19 (2.04–4.97) |
| Ischemic stroke | 10/23,999 | 0.45 (0.24–0.84) | 43/95,943 | 0.49 (0.37–0.67) | -0.04 (-0.36–0.28) | 0.92 (0.41-1.85) | 0.89 (0.44–1.77) | 0.98 (0.48–2.00) | 1.58 (0.48–5.19) | 1.01 (0.45–2.25) |
| Intracranial hemorrhage | 8/24,003 | 0.36 (0.18–0.73) | 32/95,982 | 0.37 (0.26–0.52) | 0 (-0.03–0.28) | 0.99 (0.39–2.19) | 0.95 (0.44–2.06) | 0.99 (0.44–2.20) | 0.80 (0.16–4.09) | 1.01 (0.42–2.41) |
| Pulmonary embolism | 5/24,006 | 0.23 (0.09–0.54) | 8/96,044 | 0.09 (0.05–0.02) | 0.14 (-0.07–0.34) | 2.47 (0.64–8.55) | 2.36 (0.77– | NA | NA | NA |

| | | | | | | | | | | |
|---|---|---|---|---|---|---|---|---|---|---|
| | | | | | | | 7.26) | | | |
| Major osteoporotic fractures | 49/23,842 | 2.24 (1.70–2.96) | 148/95,538 | 1.71 (1.45–2.01) | 0.53 (-0.16–1.21) | 1.31 (0.93–1.82) | 1.30 (0.94–1.81) | 1.29 (0.92–1.81) | 0.71 (0.41–1.22) | 1.30 (0.92–1.83) |
| Other fractures | 137/23,387 | 6.39 (5.40–7.55) | 425/94,442 | 4.97 (4.52–5.46) | 1.42 (0.25–2.59) | 1.29 (1.05–1.56) | 1.29 (1.06–1.56) | 1.28 (1.05–1.56) | 1.17 (0.90–1.52) | 1.28 (1.05–1.56) |
| Gastrointestinal bleeding | 157/23,676 | 7.23 (6.19–8.46) | 139/95,416 | 1.61 (1.36–1.90) | 5.63 (4.47–6.79) | 4.50 (3.56–5.70) | 4.36 (3.46–5.48) | 4.35 (3.46–5.49) | 3.60 (2.57–5.03) | 4.38 (3.46–5.53) |
| Infectious pneumonia | 559/23,262 | 26.5 (24.4–28.8) | 514/93,388 | 6.08 (5.58–6.63) | 20.4 (18.2–22.7) | 4.36 (3.86–4.92) | 4.28 (3.79–4.84) | 4.35 (3.85–4.91) | 4.66 (3.84–5.64) | 4.37 (3.86–4.95) |
| Urinary tract infection | 1,244/21,117 | 66.4 (62.8–70.2) | 2,066/85,456 | 26.9 (25.8–28.1) | 39.5 (35.7–43.4) | 2.47 (2.30–2.65) | 2.46 (2.28–2.64) | 2.45 (2.27–2.63) | 2.42 (2.17–2.69) | 2.45 (2.27–2.64) |
| Anxiety/depression | 1,105/22,257 | 55.7 (52.5–59.1) | 781/90,291 | 9.56 (8.91–10.3) | 46.2 (42.8–49.5) | 5.83 (5.31–6.39) | 5.87 (5.33–6.45) | 5.89 (5.35–6.49) | 5.57 (4.82–6.43) | 5.99 (5.42–6.61) |

CI = confidence interval, NA = not available (because the number of outcomes was too small).

*Patients with a previous history of the outcome were excluded from the analysis.

Model 1: without additional adjustment

Model 2: adjusted for hypertension, diabetes, and dyslipidemia, in addition to osteoporosis for the fracture outcome

Model 3: further adjusted for BMI, smoking history, and drinking habits as a complete case analysis

Model 4: with multiple imputation for BMI, smoking history, and drinking habits

**Supplementary Table S7. Incidence rate of each outcome in each group and its difference and ratio between the groups with and without breast cancer during 1–10 years from the index month**

| (A) Follow-up <1 year | Patients with breast cancer | | Women without breast cancer | | Rate difference (95% CI) (/1000 PY) | Rate ratio (95% CI) | Hazard ratio (95% CI) | | | |
|---|---|---|---|---|---|---|---|---|---|---|
| | No. of outcomes/ No. of patients* | Rate (95% CI) (/1000 PY) | No. of outcomes/ No. of patients* | Rate (95% CI) (/1000 PY) | | | Model 1 | Model 2 | Model 3 | Model 4 |
| Myocardial infarction | 3/19,423 | 0.05 (0.02–0.17) | 6/76,162 | 0.03 (0.01–0.06) | 0 (-0.004–0.03) | 0.84 (0.24–2.63) | 1.45 (0.34–6.24) | NA | NA | NA |
| Heart failure | 34/19,392 | 0.61 (0.43–0.85) | 58/76,106 | 0.26 (0.20–0.34) | 0.35 (0.13–0.56) | 2.31 (1.47–3.59) | 3.13 (1.93–5.08) | 3.08 (1.82–5.19) | 3.24 (1.21–8.69) | 3.34 (1.89–5.88) |
| Atrial fibrillation/flutter | 55/19,338 | 0.99 (0.76–1.29) | 158/75,935 | 0.72 (0.62–0.84) | 0.27 (-0.02–0.55) | 1.37 (0.99–1.88) | 1.37 (0.98–1.91) | 1.30 (0.92–1.83) | 1.19 (0.66–2.14) | 1.31 (0.91–1.87) |
| Ischemic stroke | 14/19,406 | 0.25 (0.15–0.42) | 84/76,057 | 0.38 (0.31–0.47) | -0.13 (-0.29–0.02) | 0.66 (0.34–1.16) | 0.61 (0.33–1.12) | 0.53 (0.28–0.99) | 0.51 (0.16–1.65) | 0.47 (0.24–0.96) |
| Intracranial hemorrhage | 11/19,411 | 0.20 (0.11–0.36) | 80/76,088 | 0.36 (0.29–0.45) | -0.17 (-0.31– -0.03) | 0.54 (0.26–1.02) | 0.51 (0.26–1.01) | 0.46 (0.23–0.93) | 0.91 (0.30–2.76) | 0.45 (0.21–0.97) |
| Pulmonary embolism | 9/19,417 | 0.16 (0.08–0.31) | 19/76,150 | 0.09 (0.05–0.14) | 0.07 (-0.04–0.19) | 1.87 (0.74–4.33) | 1.88 (0.82– | NA | NA | NA |

| | | | | | | | | | | |
|---|---|---|---|---|---|---|---|---|---|---|
| | | | | | | | 4.33) | | | |
| Major osteoporotic fractures | 190/19,255 | 3.44 (2.98–3.96) | 398/75,680 | 1.82 (1.65–2.01) | 1.62 (1.10–2.14) | 1.89 (1.58–2.25) | 1.77 (1.46–2.14) | 1.79 (1.47–2.16) | 1.94 (1.44–2.60) | 1.78 (1.46–2.17) |
| Other fractures | 506/18,819 | 9.46 (8.67–10.3) | 1,014/74,654 | 4.73 (4.44–5.03) | 4.74 (3.86–5.61) | 2.00 (1.80–2.23) | 2.07 (1.85–2.33) | 2.08 (1.86–2.34) | 2.20 (1.86–2.60) | 2.09 (1.86–2.34) |
| Gastrointestinal bleeding | 298/19,030 | 5.49 (4.90–6.15) | 378/75,577 | 1.73 (1.57–1.92) | 3.76 (3.11–4.40) | 3.17 (2.71–3.70) | 3.22 (2.73–3.79) | 3.21 (2.72–3.78) | 2.97 (2.31–3.82) | 3.22 (2.73–3.80) |
| Infectious pneumonia | 391/18,328 | 7.51 (6.81–8.30) | 968/73,677 | 4.58 (4.30–4.88) | 2.93 (2.13–3.73) | 1.64 (1.45–1.85) | 1.66 (1.46–1.88) | 1.66 (1.46–1.88) | 1.42 (1.17–1.74) | 1.67 (1.47–1.89) |
| Urinary tract infection | 1,289/16,113 | 29.3 (27.7–30.9) | 4,272/66,419 | 23.3 (22.6–24.0) | 6.02 (4.27–7.76) | 1.26 (1.18–1.34) | 1.26 (1.18–1.35) | 1.26 (1.18–1.35) | 1.20 (1.09–1.33) | 1.26 (1.18–1.35) |
| Anxiety/depression | 595/17,101 | 12.4 (11.4–13.4) | 1,730/71,220 | 8.50 (8.11–8.91) | 3.88 (2.81–4.95) | 1.46 (1.32–1.60) | 1.51 (1.36–1.67) | 1.51 (1.36–1.67) | 1.59 (1.37–1.84) | 1.51 (1.37–1.67) |

CI = confidence interval, NA = not available (because the number of outcomes was too small).

*Patients with a previous history of the outcome were excluded from the analysis.

Model 1: without additional adjustment

Model 2: adjusted for hypertension, diabetes, and dyslipidemia, in addition to osteoporosis for the fracture outcome

Model 3: further adjusted for BMI, smoking history, and drinking habits as a complete case analysis

Model 4: with multiple imputation for BMI, smoking history, and drinking habits

**Supplementary Figure S2. Additional analysis by treatment regimens**

(A) Heart failure

By chemotherapy

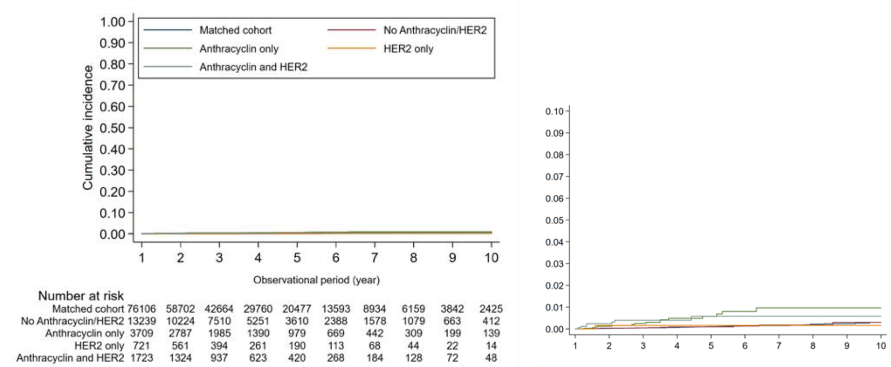

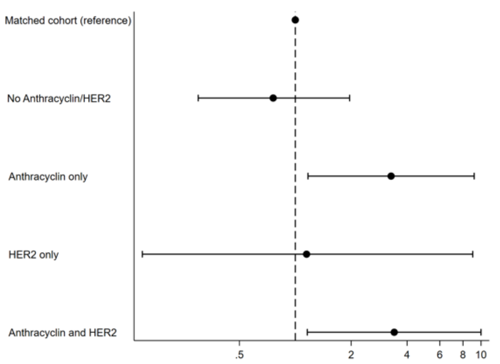

Adjusted hazard ratio (95% CI)

By hormone therapy

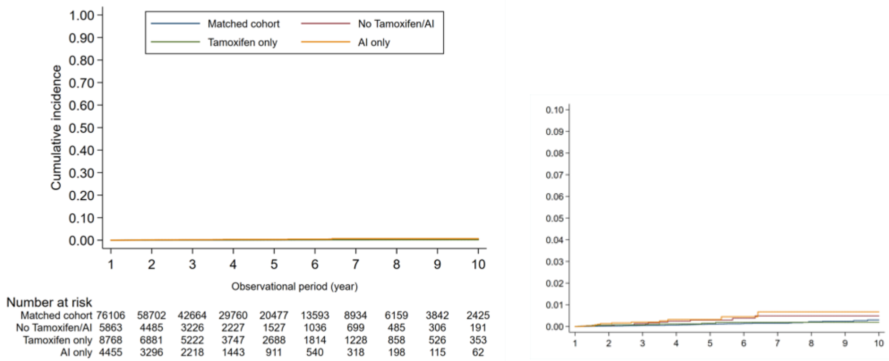

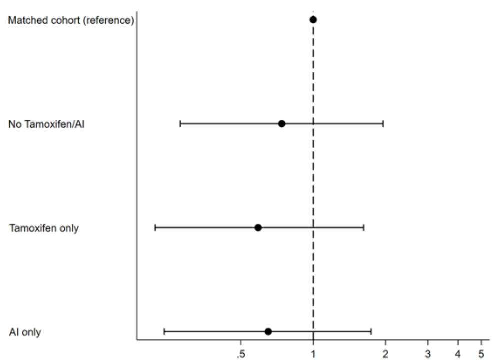

Adjusted hazard ratio (95% CI)

HER2 = human epidermal growth factor receptor 2, AI = aromatase inhibitor.

Note: 348 patients who received both tamoxifen and aromatase inhibitors within 1 year of diagnosis were excluded from the analysis.

*Adjusted for age, year of index date, hypertension, diabetes, dyslipidemia, axillary dissection, radiotherapy, tamoxifen, and aromatase inhibitors

**Adjusted for age, year of index date, hypertension, diabetes, dyslipidemia, axillary dissection, radiotherapy, anthracycline, and HER2-targeted therapy

(B) Atrial fibrillation/flutter

By chemotherapy

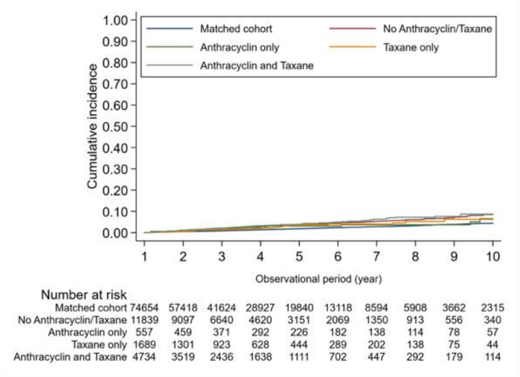

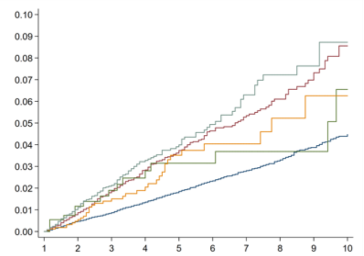

By hormone therapy

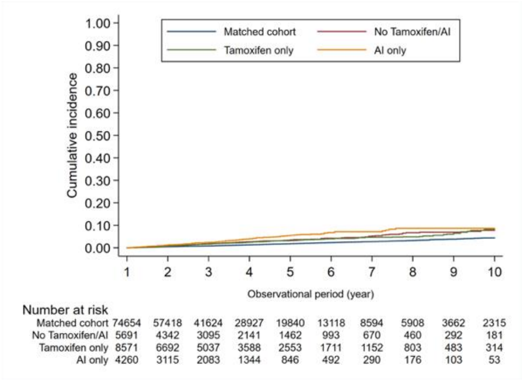

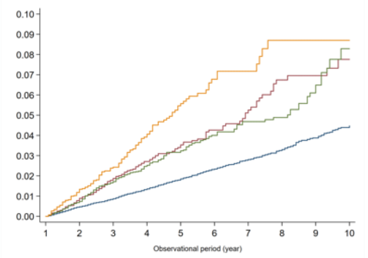

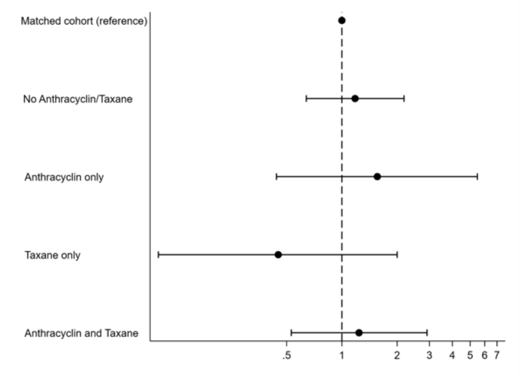

Adjusted hazard ratio (95% CI)

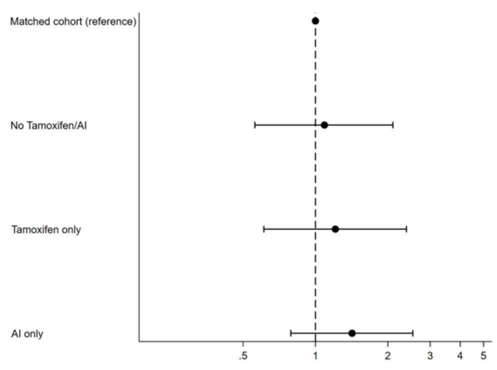

Adjusted hazard ratio (95% CI)

AI = aromatase inhibitor.

Note: 348 patients who received both tamoxifen and aromatase inhibitors within 1 year of diagnosis were excluded from the analysis.

*Adjusted for age, year of index date, hypertension, diabetes, dyslipidemia, axillary dissection, radiotherapy, tamoxifen, and aromatase inhibitors

**Adjusted for age, year of index date, hypertension, diabetes, dyslipidemia, axillary dissection, radiotherapy, anthracycline, and taxane

(C) Major osteoporotic fractures

By chemotherapy

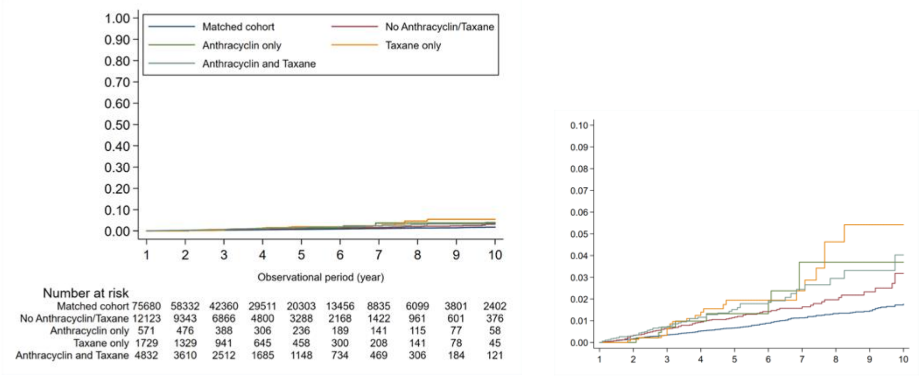

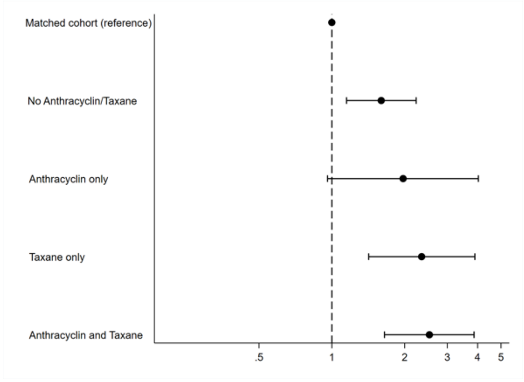

Adjusted hazard ratio (95% CI)

By hormone therapy

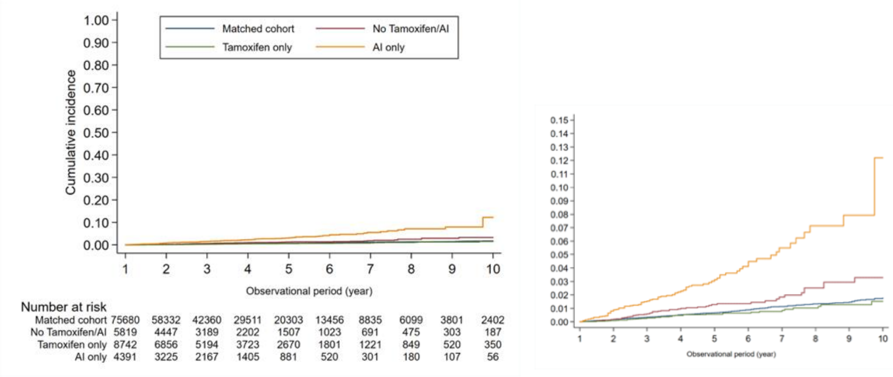

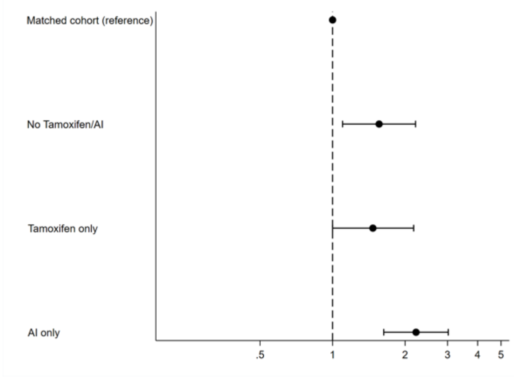

Adjusted hazard ratio (95% CI)

AI = aromatase inhibitor.

Note: 348 patients who received both tamoxifen and aromatase inhibitors within 1 year of diagnosis were excluded from the analysis.

*Adjusted for age, year of index date, hypertension, diabetes, dyslipidemia, osteoporosis, axillary dissection, radiotherapy, tamoxifen, and aromatase inhibitors

**Adjusted for age, year of index date, hypertension, diabetes, dyslipidemia, osteoporosis, axillary dissection, radiotherapy, anthracycline, and taxane

(D) Other fractures

By chemotherapy

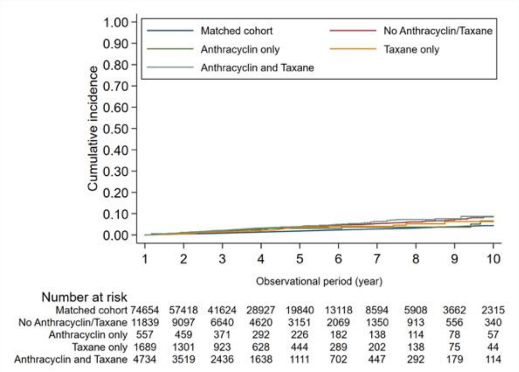

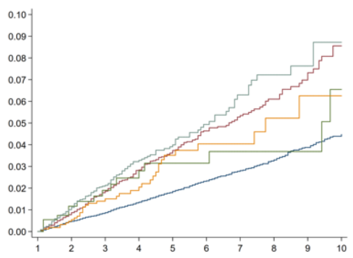

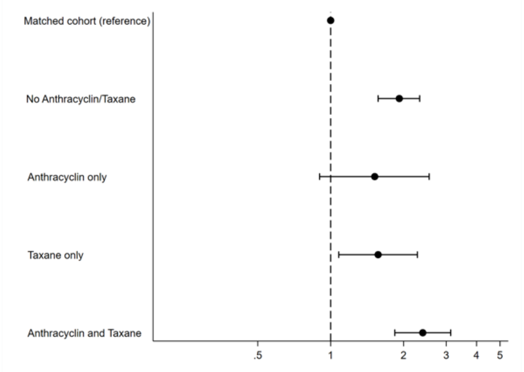

Adjusted hazard ratio (95% CI)

By hormone therapy

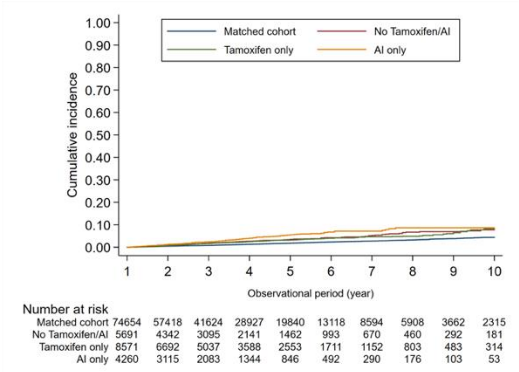

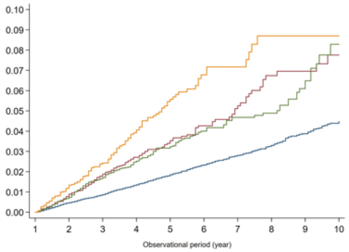

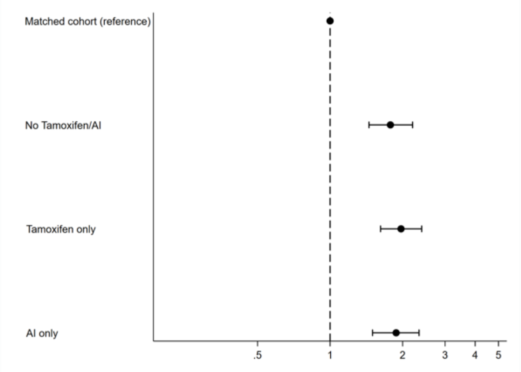

Adjusted hazard ratio (95% CI)

AI = aromatase inhibitor.

Note: 348 patients who received both tamoxifen and aromatase inhibitors within 1 year of diagnosis were excluded from the analysis.

*Adjusted for age, year of index date, hypertension, diabetes, dyslipidemia, osteoporosis, axillary dissection, radiotherapy, tamoxifen, and aromatase inhibitors

**Adjusted for age, year of index date, hypertension, diabetes, dyslipidemia, osteoporosis, axillary dissection, radiotherapy, anthracycline, and taxane

(E) Gastrointestinal bleeding

By chemotherapy

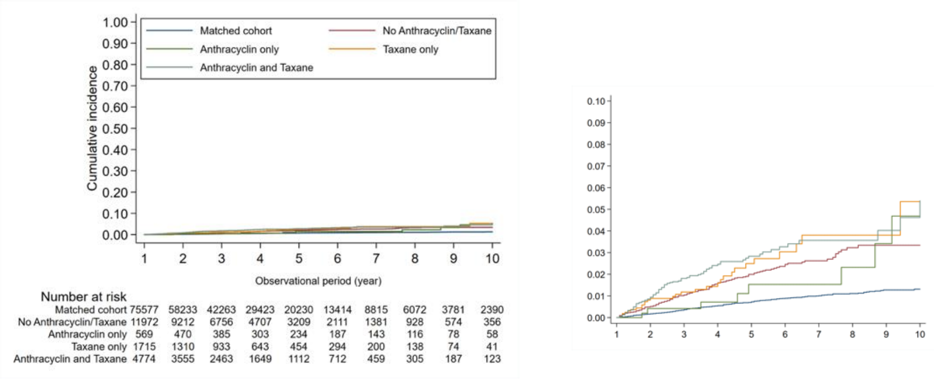

By hormone therapy

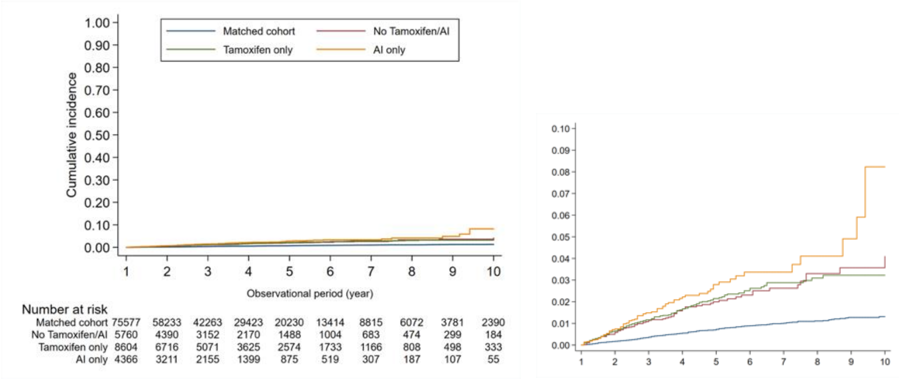

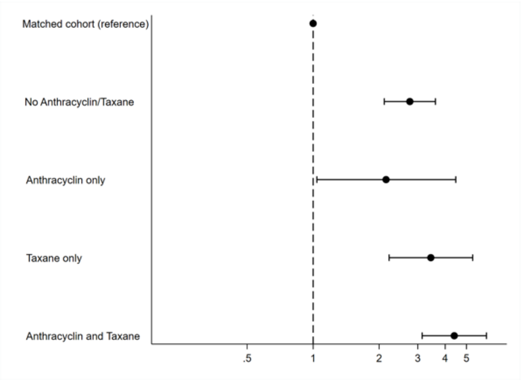

Adjusted hazard ratio (95% CI)*

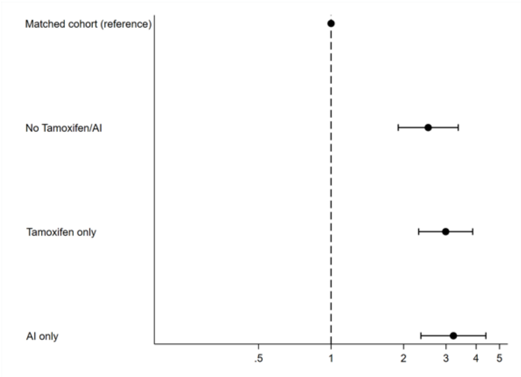

Adjusted hazard ratio (95% CI)**

AI = aromatase inhibitor.

Note: 348 patients who received both tamoxifen and aromatase inhibitors within 1 year of diagnosis were excluded from the analysis.

*Adjusted for age, year of index date, hypertension, diabetes, dyslipidemia, axillary dissection, radiotherapy, tamoxifen, and aromatase inhibitors

**Adjusted for age, year of index date, hypertension, diabetes, dyslipidemia, axillary dissection, radiotherapy, anthracycline, and taxane

(F) Infectious pneumonia

By chemotherapy

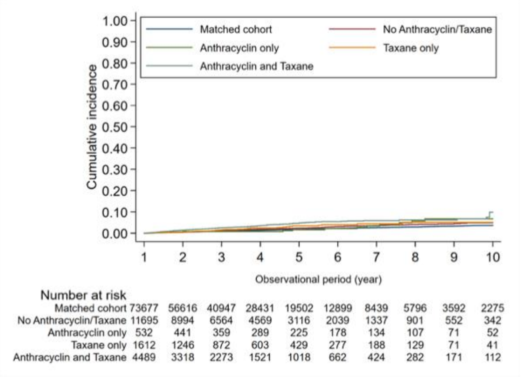

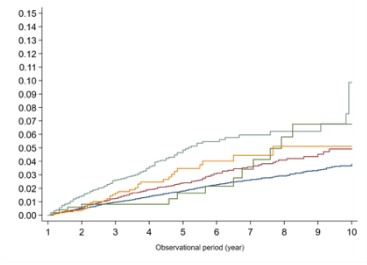

By hormone therapy

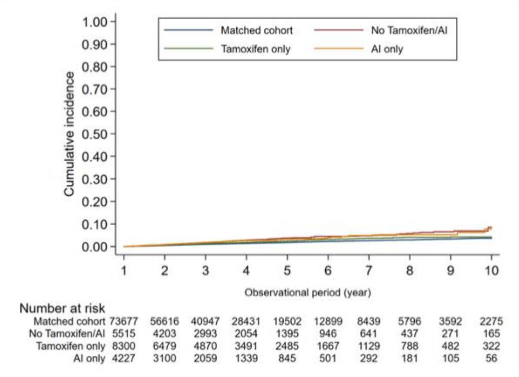

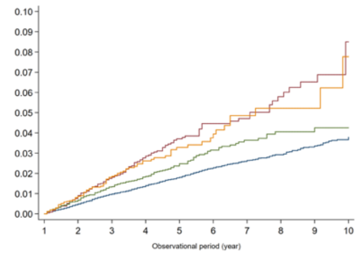

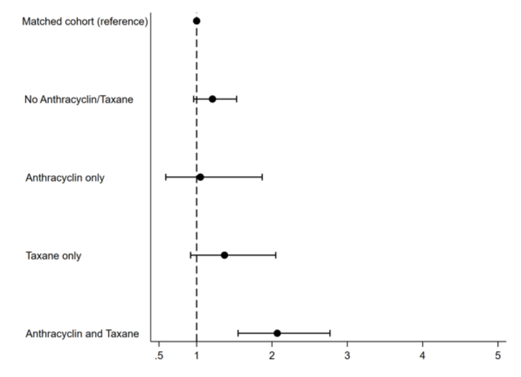

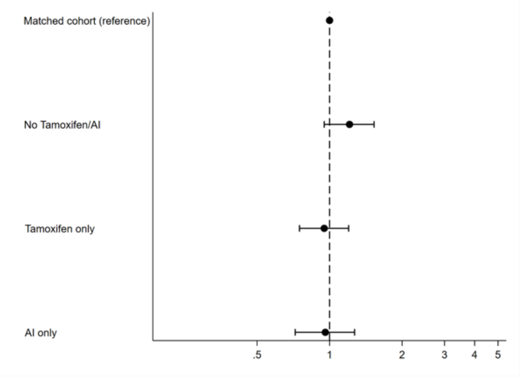

Adjusted hazard ratio (95% CI)*

Adjusted hazard ratio (95% CI)**

AI = aromatase inhibitor.

Note: 348 patients who received both tamoxifen and aromatase inhibitors within 1 year of diagnosis were excluded from the analysis.

*Adjusted for age, year of index date, hypertension, diabetes, dyslipidemia, axillary dissection, radiotherapy, tamoxifen, and aromatase inhibitors

**Adjusted for age, year of index date, hypertension, diabetes, dyslipidemia, axillary dissection, radiotherapy, anthracycline, and taxane

(G) Urinary tract infection

By chemotherapy

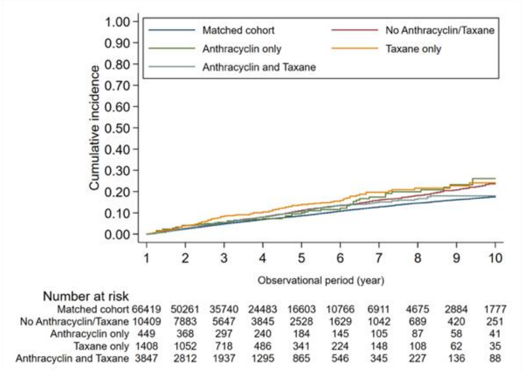

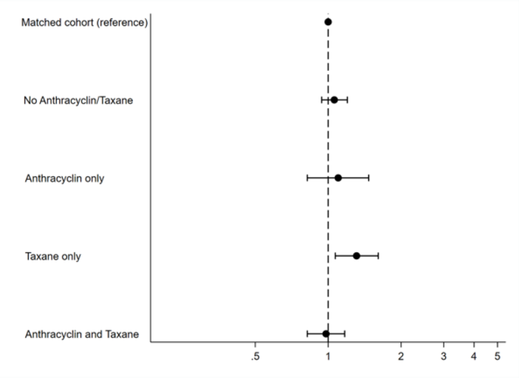

Adjusted hazard ratio (95% CI)*

By hormone therapy

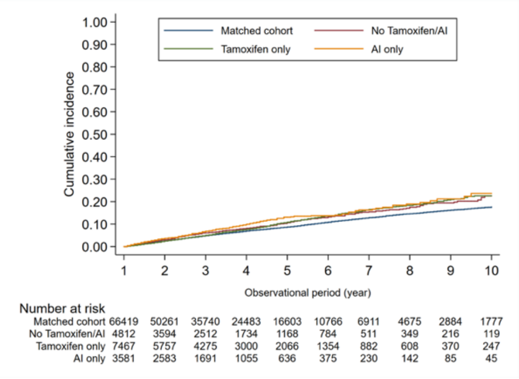

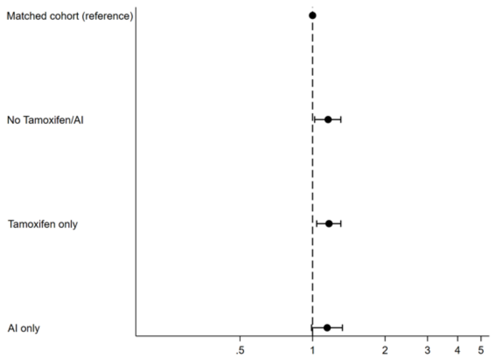

Adjusted hazard ratio (95% CI)**

AI = aromatase inhibitor.

Note: 348 patients who received both tamoxifen and aromatase inhibitors within 1 year of diagnosis were excluded from the analysis.

*Adjusted for age, year of index date, hypertension, diabetes, dyslipidemia, axillary dissection, radiotherapy, tamoxifen, and aromatase inhibitors

**Adjusted for age, year of index date, hypertension, diabetes, dyslipidemia, axillary dissection, radiotherapy, anthracycline, and taxane

(H) Anxiety/depression

By chemotherapy

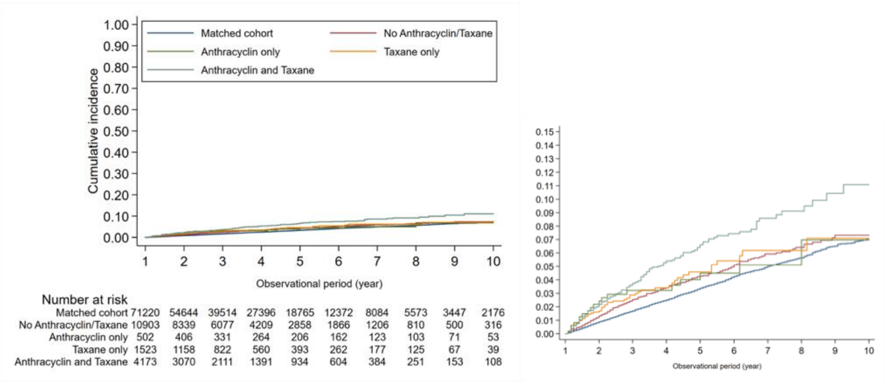

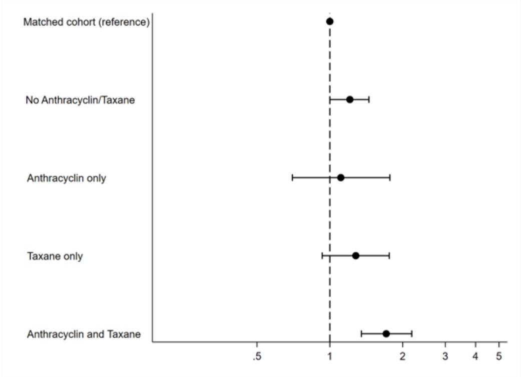

Adjusted hazard ratio (95% CI) *

By hormone therapy

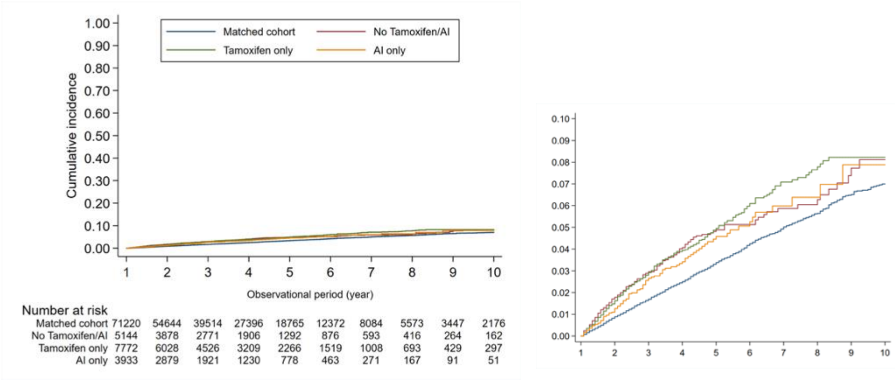

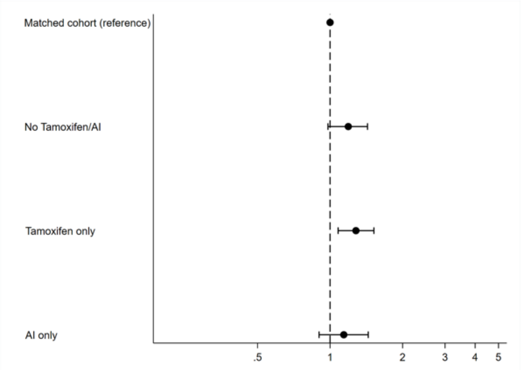

Adjusted hazard ratio (95% CI)**

AI = aromatase inhibitor.

Note: 348 patients who received both tamoxifen and aromatase inhibitors within 1 year of diagnosis were excluded from the analysis.

*Adjusted for age, year of index date, hypertension, diabetes, dyslipidemia, axillary dissection, radiotherapy, tamoxifen, and aromatase inhibitors

**Adjusted for age, year of index date, hypertension, diabetes, dyslipidemia, axillary dissection, radiotherapy, anthracycline, and taxane
